# Patterns of oncogene co-expression at single cell resolution in cancer influence survival

**DOI:** 10.1101/2020.10.20.20216101

**Authors:** Michal Marek Hoppe, Patrick Jaynes, Fan Shuangyi, Yanfen Peng, Phuong Mai Hoang, Liu Xin, Sanjay De Mel, Limei Poon, Esther Chan, Joanne Lee, Choon Kiat Ong, Tiffany Tang, Soon Thye Lim, Chandramouli Nagarajan, Nicholas Francis Grigoropoulos, Soo-Yong Tan, Susan Swee-Shan Hue, Sheng-Tsung Chang, Shih-Sung Chuang, Shaoying Li, Joseph D. Khoury, Hyungwon Choi, Carl Harris, Alessia Bottos, Laura J. Gay, Hendrik F. P. Runge, Ilias Moutsopoulos, Irina Mohorianu, Daniel J. Hodson, Pedro Farinha, Anja Mottok, David W. Scott, Gayatri Kumar, Kasthuri Kannan, Wee-Joo Chng, Yen Lin Chee, Siok-Bian Ng, Claudio Tripodo, Anand D. Jeyasekharan

## Abstract

**Background:** Cancers often overexpress multiple clinically relevant oncogenes. However, it is not known if multiple oncogenes within a cancer combine uniquely in specific cellular sub-populations to influence clinical outcome. We studied this phenomenon using the prognostically relevant oncogenes MYC, BCL2 and BCL6 in Diffuse Large B-Cell Lymphoma (DLBCL).

**Methods:** Quantitative multispectral imaging simultaneously measured oncogene co-expression at single-cell resolution in reactive lymphoid tissue (*n*=12) and four independent cohorts (*n*=409) of DLBCL. Mathematically derived co-expression phenotypes were evaluated in DLBCLs with immunohistochemistry (*n*=316) and eight DLBCL cohorts with gene expression data (n=4186). Bulk and single-cell RNA sequencing was performed on patient-derived B-cells with induced co-expression of MYC, BCL2 and BCL6.

**Results:** Unlike in non-malignant lymphoid tissue where the co-expression of MYC, BCL2 and BCL6 in a B-cell is limited, DLBCLs show multiple permutations of oncogenic co-expression in malignant B-cells. The percentage of cells with a unique combination MYC+BCL2+BCL6-(M+2+6-) consistently predicts survival in contrast to that of other combinations (including M+2+6+). An estimated percentage of M+2+6-cells can be derived from any quantitative measurement of the component individual oncogenes, and correlates with survival in immunohistochemistry and gene expression datasets. Comparative transcriptomic analysis of DLBCLs and transformed patient-derived B-cells identifies cyclin D2 (CCND2) as a potential BCL6-repressed regulator of proliferation in the M+2+6-population.

**Conclusions:** Unique patterns of oncogene co-expression at single-cell resolution affect clinical outcomes in DLBCL. Similar analyses evaluating oncogenic combinations at the cellular level may impact diagnostics and target discovery in other cancers.

## Introduction

Oncogene overexpression is common in cancer. The concomitant increase in oncogenic proteins (oncoproteins) influences both prognosis and treatment^1^. Notable examples routinely assessed in clinical practice include HER2 in breast cancer and ALK in lung cancer. However, cancers often overexpress more than one oncogene. Whether multiple oncogenes interact at the single-cell level to influence clinical outcome remains an important unresolved question. This is particularly relevant since cancers are a heterogenous mosaic of tumor cell sub-populations^2^, and oncogenes show clinically significant intratumor heterogeneity (ITH) in expression^1^. While only a subset of cells within a tumor co-express all oncogenes measured at the whole-tumor scale, clinical techniques for oncogene estimation in cancer (such as immunohistochemistry, IHC) study them in isolation. It is therefore still not known if subsets of cells within a given cancer expressing specific combinations of oncogene drive eventual clinical phenotypes.

We aimed to address this question using multiplexed fluorescent immunohistochemistry (mfIHC), a technique that can simultaneously and quantitatively evaluate a set of proteins with single-cell resolution. This allows measurement of single-cell oncogene co-expression from sufficient samples for robust multivariate correlations with clinical outcome. We chose Diffuse Large B-cell Lymphoma (DLBCL) as a model to evaluate the clinical impact of ITH in oncogene co-expression. DLBCL is the most common aggressive lymphoma worldwide^3^, and overexpression of the oncogenes MYC, BCL2 and BCL6 influence pathogenesis and prognosis^4-6^. However, there is significant variability among studies regarding the prognostic significance of these oncogenes with debate on appropriate cut-off thresholds to define “positivity”. These considerations offer an ideal scenario to evaluate whether these oncogenes show differential co-expression at the single cell level in DLBCL, and to investigate how they collaborate or influence each other at the cellular level to affect survival.

## Methods

### Samples and datasets

Non-malignant samples included chronic tonsillitis (*n*=10) and reactive lymph nodes (*n*=2) collected at the National University Hospital (NUH) Singapore (DSRB protocol 2015/00176). DLBCL samples in tissue microarray (TMA) format were from NUH (*n*=152), Chi-Mei Medical Center, Taiwan (CMMC; *n*=150), Singapore General Hospital, Singapore (SGH; *n*=67) and the MD Anderson Cancer Center, USA (MDA; *n*=40), and the British Columbia Cancer Agency (BCA; *n*=274) - used for validation^7^. Datasets GSE117556 (*n*=928)^8^, GSE125966 (*n*=553), GSE31312 (*n*=498)^9^, GSE10846 (*n*=420)^10^, GSE32918 (*n*=140)^11^ and GSE87371 (*n*=221)^12^ were obtained from the Gene Expression Omnibus (GEO), while those from Reddy *et al* (*n*=773)^13^ and Schmitz *et al* (*n*=480)^14^ were from the database of Genotypes and Phenotypes (dbGaP). The GOYA dataset (GSE125966) was analysed independently by F. Hoffmann-La Roche Ltd.

### Quantitative multiplexed fluorescent immunohistochemistry (mfIHC) and scoring

Quantitative multiplexed fluorescent immunohistochemistry (mfIHC) was performed using sequential OPAL-TSA staining as described previously^15^ (Supplementary table1 and Supplementary table2). Images were acquired using the Vectra imager and analysed using inForm2.4.8 (Akoya Biosciences, USA). DAPI nuclear staining and CD20 membrane staining were used to segment cells. A pathologist-evaluated fluorescent intensity threshold defined “positive” and “negative” cells for CD20, MYC, BCL2 and BCL6 per tissue microarray (TMA) core. For every cell in a given case, information on CD20, MYC, BCL2 and BCL6 allowed categorization into one of the following permutations: M+2+6+, M+2+6-, M+2-6+, M+2-6-, M-2+6+, M-2+6-, M-2-6+ and M-2-6- (where M is MYC, 2 is BCL2, and 6 is BCL6 and + or – indicates the presence or absence of the oncogene) within the CD20+ B-cell compartment.

### Survival analysis

Where used, all variables satisfied proportional hazards assumptions for Cox proportional hazard (Cox PH) models. Hazard ratios are displayed per unit (in 5% increments) of percentages of subpopulations as continuous variables. For Kaplan-Meier analyses, the cohorts were dichotomized at an optimal cut-off. Statistical tests were two-sided and *p*≤0.05 was considered as statistically significant. Statistical tests were performed using SPSS23, GraphPad Prism9 and R.

### Supplementary methods/ table

The supplementary methods contains details on patient characteristics, probabilistic subpopulation percentage prediction, transformation of RNA data, semi-quantitative IHC, mutational and gene expression analysis, and manipulation of tonsil-derived GC B cells.

### Data Sharing Statement

RNAseq data is deposited to GEO^16^ (https://www.ncbi.nlm.nih.gov/geo/) GSE203446. Requests for primary image data/protocols can be made to Dr Anand Jeyasekharan via.

## Results

### Physiological patterns of MYC, BCL2 and BCL6 co-expression are disrupted in DLBCL

We first compared the co-expression of the oncogenes MYC, BCL2 and BCL6 between reactive lymphoid tissue and DLBCL (Supplementary figure1A). Consistent with known physiology, BCL6 and BCL2 expression was restricted to reactive lymphoid germinal centres (GC) and extra-GC regions respectively, while MYC showed sparse positivity in the GC CD20+ cells^17^ (Figure1A, Supplementary figure1B). A binary +/- map for each oncogene facilitates the quantitation of “sub-populations” of cells based on MYC BCL2 and BCL6 co-expression (Supplementary figure1C). The repertoire of sub-populations defined by MYC/BCL2/BCL6 in reactive lymphoid B-cells was limited, with M-2-6+ being dominant within the GC, and M-2+6-being dominant outside the GC (Figure1B). Hardly any cells displayed co-expression of all three oncogenes MYC, BCL2 and BCL6 in either compartment, consistent across several reactive lymphoid tissues analysed (Figure1C, Supplementary figure1D).

**Figure1.**
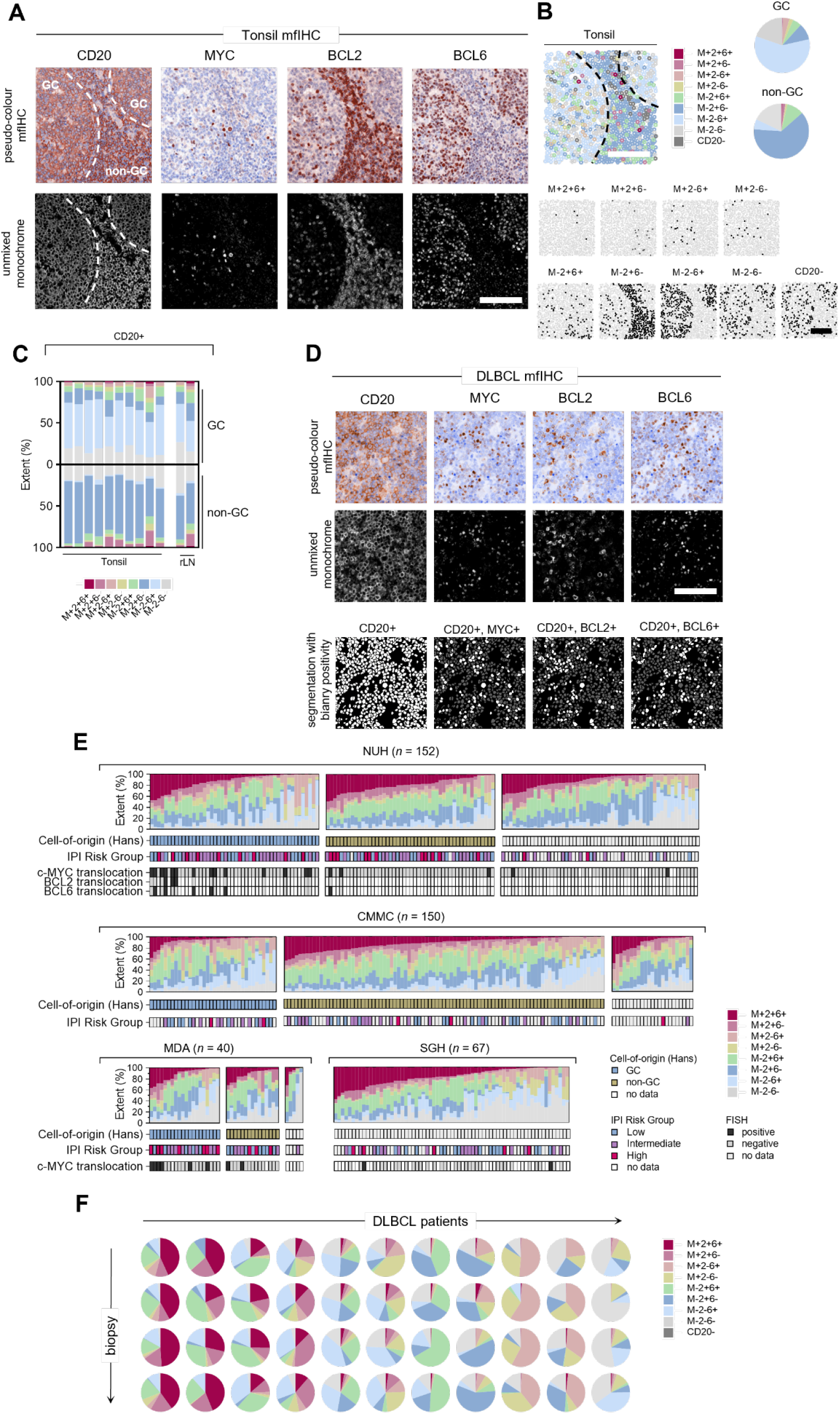
Quantitative single-cell analysis of MYC, BCL2 and BCL6 protein expression in B-cells in normal tissues and diffuse large B-cell lymphoma. A, Spectrally unmixed multiplexed fluorescent images for CD20, MYC, BCL2 and BCL6 and nuclear counterstaining in tonsil tissue. The germinal center (GC) and non-germinal center (non-GC) zones are indicated. Scale bar is 100μm. Marker-positivity is indicated. B, Spatial map of MYC/BCL2/BCL6 sub-populations, i.e. possible permutations of MYC/BCL2/BCL6-positivity and -negativity within the CD20-positive cell population in a tonsil image. Scale bars are 100μm. C, Quantitation of sub-population extent within CD20-positive cells in normal samples resolved spatially between the GC and non-GC zones. D, Example of MYC/BCL2/BCL6/CD20 multiplexed fluorescent immunohistochemistry (mfIHC) staining in diffuse large B-cell lymphoma (DLBCL). Spectrally unmixed false-coloured and monochrome images for CD20, MYC, BCL2 and BCL6 are shown (top and middle panels). Cell-segmentation and single-marker positivity masks are shown within the CD20-positive cell population (bottom panel). Scale bar is 100μm. E, Summary of percentage extent of sub-populations across patients from National University Hospital (NUH), Chi-Mei Medical Center (CMMC), MD Anderson (MDA) and Singapore General Hospital (SGH). Relevant clinicopathological features are indicated, see also Supplementary figure2. Patients are grouped according to Cell-of-origin based on the Hans algorithm and sorted arbitrarily according to extent of the triple-positive M+2+6+ sub-population extent. IPI Risk Group - International Prognostic Index Risk Group, GC - germinal center B-cell like lymphoma. FISH - fluorescence in situ hybridization. F, Intra-patient spatial stability of sub-populations - proportion of sub-populations measured across four spatially distinct biopsies from the same patient (rows). Biopsy comparison overview is shown across eleven example DLBCL patients (columns).

In contrast, DLBCL cells frequently co-express these three oncogenes (Figure1D, Supplementary figure1E, and Supplementary figure2). Even within cases characterized by high overall levels of MYC, BCL2 and BCL6 expression, these three oncogenes were not always found in the same cells, underscoring intratumoral heterogeneity (ITH) in DLBCL. The percentage of each sub-population was variable between patients but was remarkably similar in overall distribution and clustering among four cohorts (Figure1E, Supplementary figure2). The percentages of sub-populations were stable across distinct TMA cores from the same patient, confirming them to show patient-specific profiles (Figure1F, Supplementary figure1F). These sub-populations were not associated with clinico-pathological features such as age, gender, International Prognostic Index (IPI) Risk Group, cell-of-origin (COO, IHC Hans algorithm^18^), nor were they associated with MYC/BCL2/BCL6 translocation status (Supplementary figure3) - confirming previous observations that translocations do not account for the majority of MYC, BCL2 and BCL6 overexpression in DLBCL^19^. Pair correlation function (PCF) analyses on single-cell resolved image data with spatial coordinates^20^ show that subpopulations cluster in space and do not display a random (Poisson) distribution (Supplementary figure4 and Supplementary Results), in keeping with local ecosystems for proliferation and evolution within a growing tumor.

### Cells co-expressing MYC and BCL2 without BCL6 confer poor survival in DLBCL

We next evaluated the relationship between MYC/BCL2/BCL6 sub-populations and prognosis, using pretreatment biopsies of R-CHOP treated DLBCL cases from 3 cohorts (NUH, *n*=98, SGH, *n*=41; MDA, *n=*36). To avoid arbitrary cut-offs, we evaluated the percentage of cells with oncogenic combinations as a continuous variable in a univariate cox proportional hazards (Cox PH) analysis for overall survival (OS). Though there was expected variability between cohorts in the prognostic impact of MYC, BCL2 and BCL6 as individual oncogenes^21^, the percentage of M+2+6- cells stood out for consistent poor prognostic significance (Figure2). This association of high M+2+6- percentage and poor outcome remained statistically significant in a multivariate Cox PH model adjusted for clinically relevant DLBCL clinicopathological parameters of IPI Risk Group, COO and MYC FISH status (Supplementary table3; Supplementary table4). These results suggest that the prognostic impact of these oncogenes in DLBCL is driven by a unique sub-population of cells expressing MYC and BCL2 without BCL6.

### A probabilistic model accurately predicts MYC, BCL2 and BCL6 co-expression in DLBCL

As the percentage of MYC+BCL2+BCL6- (M+2+6-) cells correlates with poor survival, we wanted to check if this percentage could be inferred mathematically. We first evaluated the relationship between these oncogenes in DLBCL at the single cell-level (Figure3A). In contrast to mutually exclusive expression within specific topological compartments in reactive lymphoid tissues, MYC, BCL2 and BCL6 do not show any correlation with each other in DLBCL (Figure3B). This suggests that independent gene regulatory mechanisms drive expression of these oncogenes in DLBCL, and that cellular co-expression therefore is a stochastic outcome (i.e. based on probability alone). An implication of this phenomenon is that the percentage of any sub-population are predictable using a probabilistic model, if the percentages of each component oncogene are known. In this model, the percentage of any given subpopulation is derived by multiplying proportions for presence or absence of each individual marker comprising the sub-population (Supplementary Methods). We validated this hypothesis using our single-cell resolved mfIHC data, observing a highly concordant correlation between observed and predicted percentages for each sub-population (Figure3C, Supplementary figure5).

**Figure2.**
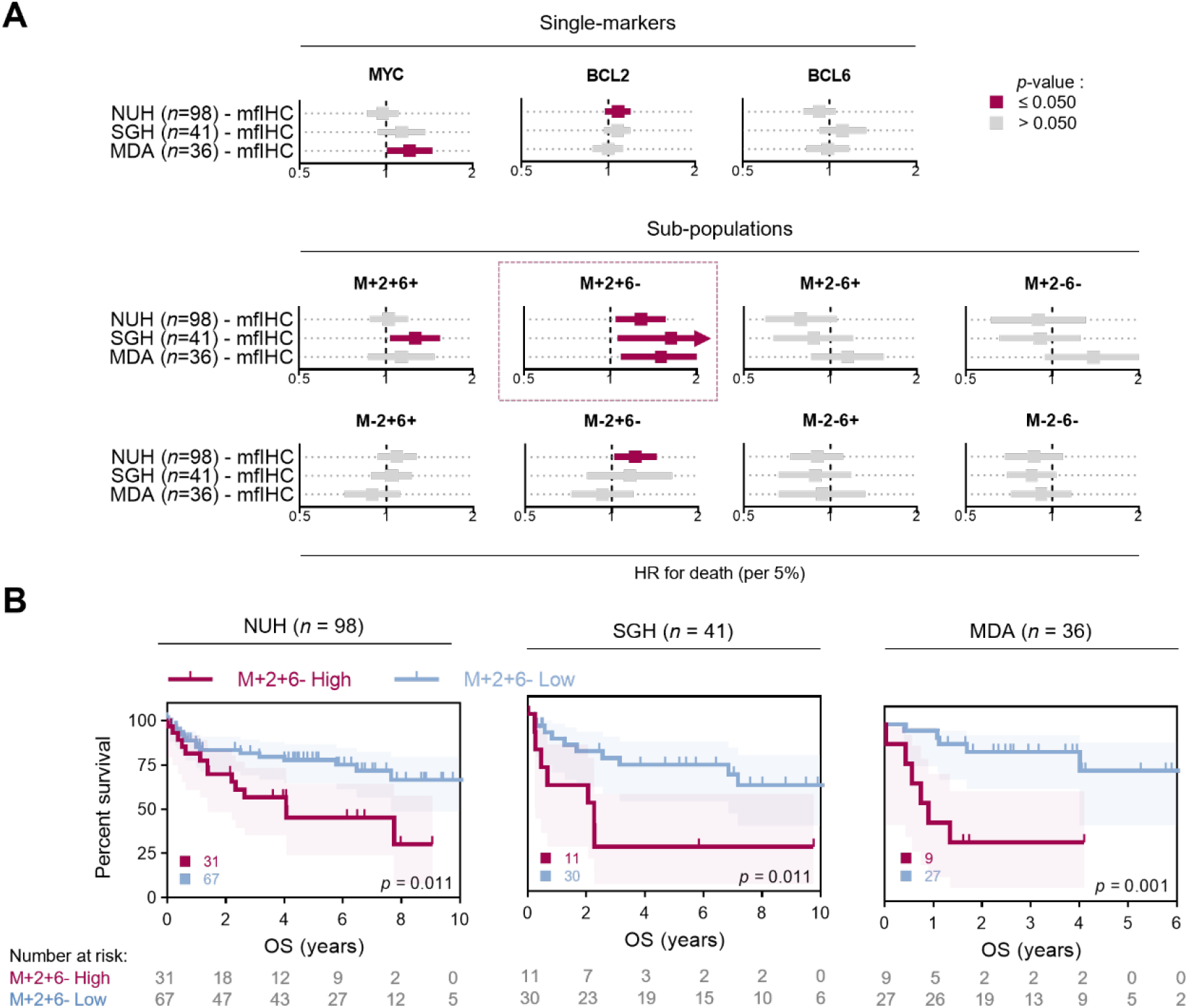
Prognostic significance of sub-populations after R-CHOP therapy. A, Univariate Cox proportional hazards model analysis for MYC, BCL2 and BCL6 single-marker and sub-populations percentage extent in the NUH, SGH and MDA cohorts. Percentage-extent was used as a continuous variable in the model at 5% increments (see Methods). OS – overall survival. Hazard ratio (HR) with 95% confidence interval per 5%-positivity increment is shown. B, Kaplan-Meier overall survival analysis of the NUH, SGH and MDA cohorts stratified into M+2+6- High and Low groups. Log-rank test, shading denotes 95% confidence interval. An optimal dichotomization cut-off was used for stratification; total patient numbers in each group are shown.

**Figure3.**
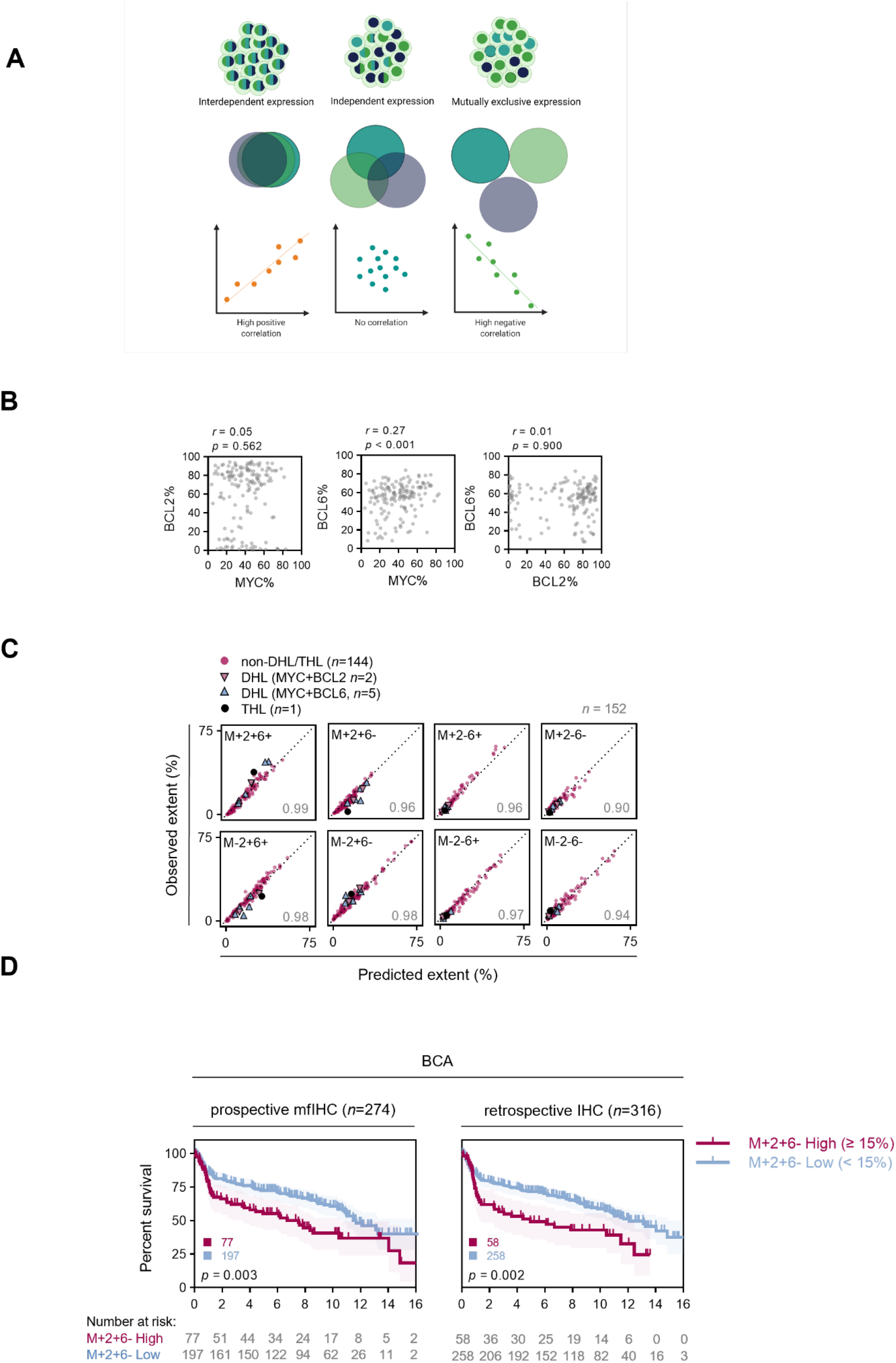
MYC, BCL2 and BCL6 protein expression is independently distributed in B-cells and therefore can be inferred form quantitative data. A, Schematic of possible relationships between expression of three oncogenes in a population. The distribution of these oncogenes can either reflect interdependent expression, independent/stochastic expression, or mutually exclusive expression. These relationships result in the percentage extent of oncogenes in the tumor being either strongly positive correlated, not correlated or strongly negatively correlated, respectively. B, Correlation of MYC, BCL2 and BCL6 percentage extent in the NUH cohort. Spearman correlation. C, Good correlation between predicted sub-population percentage extent based on single-marker positivity and observed percentage extent in the NUH cohort. Cases of double-hit lymphoma (DHL, MYC+BCL2+ translocations or MYC+BCL6+ translocations) or triple-hit lymphoma (THL) are highlighted. Spearman rho for each correlation is shown. Similar graphs for the CMMC, SGH and MDA cohorts can be found in Supplementary figure5. D, Kaplan-Meier overall survival analysis of the BCA cohort stratified into M+2+6- High and Low groups across an absolute value of 15 percentage extent positivity in prospective mfIHC validation (*left*) and retrospective IHC images (*right*). Log-rank test, shading denotes 95% confidence interval.

An extension of these findings is that any quantitative data on MYC, BCL2 and BCL6 allows estimation of the percentage of each sub-population characterized by their co-expression. Visual scoring of MYC, BCL2 and BCL6 percentage on chromogenic IHC remains the reference method for assessment of these oncogenes in clinical practice. We checked if our algorithm could estimate prognostic MYC, BCL2, BLC6 sub-populations from clinical-grade pathologist scores for MYC, BCL2 and BCL6 chromogenic IHC in a well-characterized cohort of DLBCL^22^. High M+2+6- percentage, whether directly measured by mfIHC or inferred from visual chromogenic IHC scores for single oncogenes, remained prognostic in this cohort (Figure3D), confirming the applicability of this algorithm to clinical IHC scoring in DLBCL.

### Estimation of MYC, BCL2 and BCL6 co-expressing sub-populations can be extended to gene expression data

We hypothesized that if the percentage of MYC/BCL2/BCL6 sub-populations could be inferred from individual oncogene components on IHC, they can be inferred from any quantitative data measuring MYC/BCL2/BCL6 expression-including gene expression profiling (GEP)^23^. mRNA levels of MYC, BCL2 and BCL6 do not correlate with each other (Figure4A-B), confirming their independent regulation at the transcriptional level as well. Remarkably, across eight distinct cohorts of DLBCL patients with GEP data, mRNA-inferred M+2+6- percentage proved again to be the only subpopulation consistently associated with poor survival (Figure4C; Supplementary Methods, Supplementary figure6 and Supplementary table5). This was further corroborated in a multivariate Cox PH analysis correcting for IPI Risk Group and COO (where the data was available; Supplementary table6). This GEP-focused analysis of M+2+6- was further independently replicated and validated in the GOYA clinical trial cohort^24^ (Supplementary figure7). Using these large GEP datasets, we establish an optimal dichotomization cut-off for M+2+6- at ≥15% to reliably stratify M+2+6- high patients for poor prognosis (Figure4D, Supplementary table7).

**Figure4.**
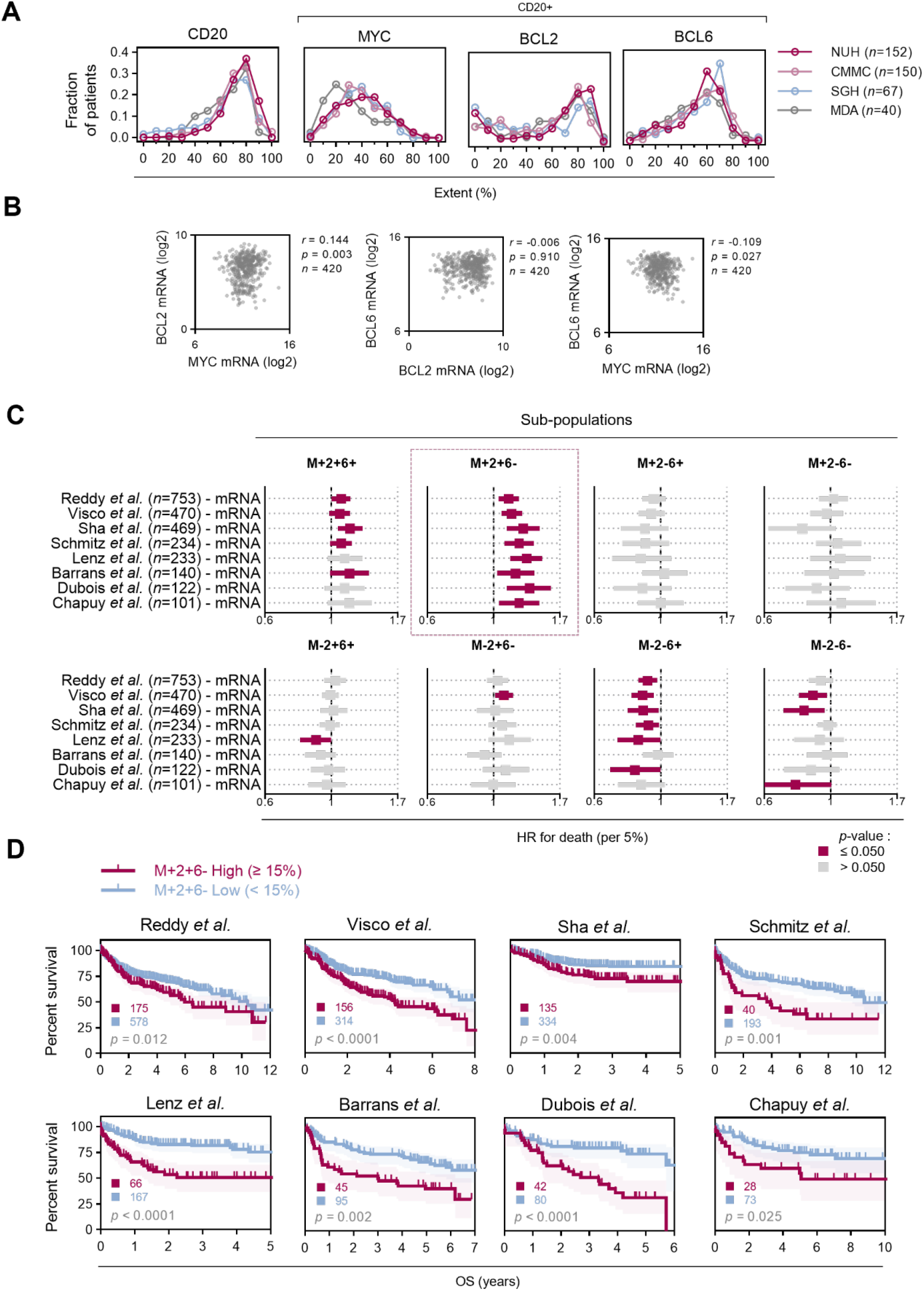
Validation of prognostic significance of inferred sub-populations extent after R-CHOP therapy in GEP datasets. A, Distribution of single-marker positivity in four DLBCL cohorts as assessed by mfIHC (See also Supplementary figure8A). B, Correlation of MYC, BCL2 and BCL6 mRNA expression in the Lenz *et al*. cohort. Spearman correlation. C, Impact of inferred MYC, BCL2 and BCL6 sub-populations in gene-expression profiling (GEP) datasets. Univariate Cox proportional hazards model analysis; percentage-extent was used as a continuous variable in the model at 5% increments. D, Kaplan-Meier overall survival analysis of GEP cohorts stratified uniformly across absolute 15% M+2+6- subpopulation extent into -High and -Low groups. Log-rank test, shading denotes 95% confidence interval. Total patient numbers in each group are shown.

### *CCND2* is enriched in the M+2+6- subpopulation and enhances its proliferative potential

Inference of oncogene co-expression from GEP datasets allows a comparative analysis with other molecular characteristics in DLBCL. Inferred M+2+6- percentage extent was consistently associated with the unfavourable MCD and A53 genetic subtypes of DLBCL (Figure5A)^25^. We also compared the inferred M+2+6- sub-population percentage with the expression of 15314 consensus genes across 3873 patients (Figure5B, Supplementary table8). Sixty-seven genes consistently correlated either positively or negatively with M+2+6- percentage extent (Figure5C), the biological significance of which will require detailed study in the future. To narrow down in the first instance, we leveraged on the observation that M+2+6- percentages robustly predict for survival while M+2+6+ percentages do not. We compared gene expression of DLBCL with gene expression of primary human tonsil-derived GC B-cells immortalised by either the overexpression of MYC and BCL2 (M+2+6-) or MYC, BCL2 and BCL6 (M+2+6+)^26^. Since BCL6 is a transcriptional repressor^27^, we hypothesised that the absence of BCL6 could influence the transcriptional profile of the M+2+6- subpopulation (Figure5D, Supplementary table9). Cross-comparing genes enriched in the M+2+6- GC B cells with genes correlated with the M+2+6- population from clinical GEP datasets, we found that *CCND2* was highly enriched in both groups (Figure5E). Single-cell RNA sequencing of a tonsil-derived GC B-cell sample clearly demonstrated an inverse correlation between *CCND2* and *BCL6* expression (Figure5F), confirming prior observations that CCND2 is transcriptionally repressed by BCL6^28,29^. We then transduced *CCND2* in M+2+6+ immortalized B cells, which were characterized by low background levels of CCND2 expression (Figure5G). The transduced M+2+6+/CCND2^High^ population started as a relatively small fraction, but rapidly expanded over time eventually outgrowing the M+2+6+/CCND2^Low^ population (Figure5H), confirming that increased CCND2 can confer a fitness advantage to cells with MYC and BCL2. Cyclin D2 expression has been reported as a marker for an adverse outcome in DLBCL^18,30^. These results may explain, in part, why DLBCL patients with a high percentage extent of the M+2+6- have a poorer prognosis, although other contributing genes and cell-extrinsic features will need evaluation in-vivo. Of general significance however, these results illustrate how estimation of oncogene co-expression phenotypes through gene expression data may uncover novel biological insight.

**Figure5.**
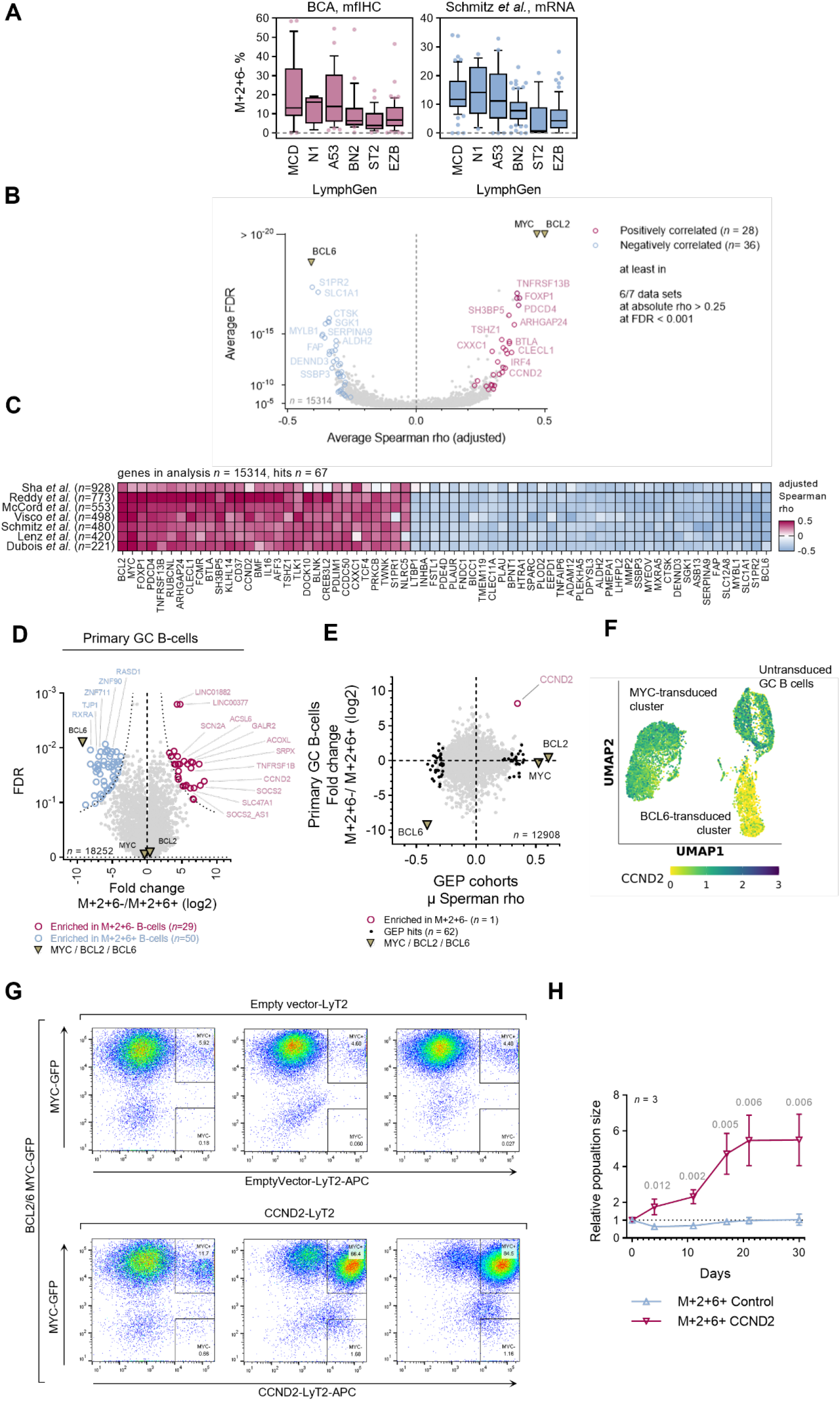
Molecular features of M+2+6- High cases and derivation and validation of cyclin D2 oncogene. A, Correlation of the M+2+6- percentage extent evaluated by mfIHC and mRNA inference with genetic subtypes (LymphGen classification). B, Volcano plot of direct correlation of gene mRNA expression and inferred M+2+6- percentage extent across seven GEP cohorts. Genes highly correlated with M+2+6- percentage extent in at least 6 out of 7 GEP cohorts at absolute Spearman rho ≥0.25 and FDR ≤0.001 are show. C, Sixty-seven hits from panel (B) criteria are shown, full analysis is available in Supplementary table8. D, Differential gene-expression between primary germinal center (GC) B-cells overexpressing M+2+ and M+2+6+. Full analysis available in Supplementary table9. E, Genes highly enriched in M+2+6- cells - correlation of results from panel (B) and (D). F, Single-cell RNA-seq of GC primary B-cells transduced either with MYC or BCL6. Expression of CCND2 is indicated in colour. G. Representative FACS plots documenting to the expansion over time of the CCND2 positive GC B-cell population in CCND2 overexpressing GC B cells (CCND2-LyT2) and non-CCND2 overexpressing GC B-cells (Empty vector LyT2). All GC B cells co-overexpress BCL2, BCL6, and MYC-GFP. H, Proliferation analysis of primary GC B-cells overexpressing M+2+6+ and CCND2.

## Discussion

In this paper, we show for the first time that sub-populations of tumor cells expressing <u>combinations</u> of oncogenes influence patient prognosis. We also show that (under conditions of independently regulated expression) these sub-populations can be inferred from quantitative single oncogene data, with a remarkable concordance to actual observed single-cell co-expression on multiplex IHC. We show two applications of predicting oncogenic sub-populations in the setting of DLBCL. First, the M+2+6- sub-population can be predicted from clinical chromogenic IHC scores, offering a refined method for utilizing MYC, BCL2 and BCL6 expression for prognostic use in DLBCL. Secondly, by estimating subpopulation percentages from GEP datasets, we demonstrate feasibility of identifying molecular features associated with a poor prognostic oncogene combination from the multitude of gene expression studies available for a given disease. Such features could identify therapeutic targets, or offer biological insight- as with our demonstration that cell cycle regulator CyclinD2 (*CCND2)* may play a role in the aggressive phenotype of M+2+6- cells. CyclinD2 promotes the G1/S transition of haematopoietic cells^31^, enhances cytokine induced-proliferation^32^, and is stabilized by EBV infection^33^, highlighting the rationale for further studies of *CCND2* in DLBCL pathogenesis and evolution.

Our single-cell resolved quantitative imaging confirms that ITH in co-expression of MYC, BCL2 and BCL6 exists in almost every case of DLBCL. This co-expression shows distinct spatial organization with non-random clustered patterns, suggesting that forces beyond genetic heterogeneity shape DLBCL evolution. These findings also suggest that quantitative assessment of the M+2+6- sub-population refines the MYC-BCL2 “double expressor lymphoma” (DEL)^4,34,35^ classification. As prior DEL classifications do not take DLBCL ITH^36,37^ into account, it was not known if DELs represent two distinct and co-existing clonal phenotypes within a lymphoma-one expressing MYC and the other BCL2. Nor was it understood why the poor outcome of DEL is exacerbated when BCL6 expression is absent^6,38^. These issues are addressed by the description of the M+2+6- subpopulation, evaluating the phenomenon at single-cell resolution. DEL remains relevant in the era of genetic DLBCL classification^39^ and novel targeted therapies. For example-patients with the DEL phenotype show improved survival on the Polatuzumab arm in comparison to the R-CHOP arm of the POLARIX trial^40^. Additional studies are required to clarify the relevance of the M+2+6- population in this setting.

This is a proof-of-concept study with several limitations. The prognostic impact of M+2+6- percentages derive from retrospective analyses and need prospective validation, along with the development of a standardized cut-off for the M+2+6- percentage. Our initial analyses from large GEP datasets suggest that >15% M+2+6- percentage extent may be a suitable starting point that is discriminatory for survival. The model we describe for prediction of oncogenic co-expression only holds true when the expression of the oncogenes are independently regulated, and will need validation in the setting of other oncogenes/cancers. Nonetheless, our demonstration that the M+2+6- phenotype confers poor survival in four empirically evaluated cohorts (mfIHC) and eight inferred cohorts (GEP) of DLBCL underscore the clinical importance of evaluating the ITH generated by the co-expression of oncogenes, and suggest that similar studies in other cancer types will be informative. Oncogene ITH occurs at multiple molecular levels in cancer: genetic^41,42^, epigenetic, transcriptomic and proteomic^43^, and affects clinical phenotypes^1,44^. Single-cell approaches to evaluate genetic^45^, transcriptomic^46,47^ and proteomic ITH have provided valuable insight into microevolutionary processes operating in cancer^48,49^. However, due to high experimental costs, the number of patients represented in scRNAseq and mass cytometry datasets are invariably small, thus precluding clinically meaningful multivariate analyses. Here we demonstrate that multiplexed microscopy through mfIHC, though limited in multiplexing potential compared to scRNAseq, is well suited to measure the clinical impact of ITH in large patient cohorts.

## Data Availability

RNAseq data is deposited to GEO (https://www.ncbi.nlm.nih.gov/geo/) under the access number: GSE203446. Requests for primary image data/protocols can be made to Dr Anand Jeyasekharan via.

https://www.ncbi.nlm.nih.gov/geo/query/acc.cgi?acc=GSE203446

## Acknowledgements

The authors acknowledge a Yong Siew Yoon Research Grant to ADJ from the National University Cancer Institute of Singapore towards the purchase of the Vectra 2 multispectral imaging system microscope.

## Authorship Contributions

Conceptualization-MMH, ADJ

Methodology (Experimental and analytical design)- MMH, FS, HC, WJC, SBN, CT, ADJ

Project administration (Coordination of sample provision/ethics approvals) - LX, SDM, LP, AB

Investigation (Quantitative mfIHC staining/ imaging) FS, YP, PMH

Data curation (Collection and curation of clinical data): LX, SDM, LP, EC, JL, YLC, DWS, OCK, TT, LST, CN, NFG, JDK

Formal analysis (Pathological review of cases): SYT, SSSH, STC, SSC, SBN, PF, AM, SL

Formal analysis (Multiplex analysis) MMH, YP, FS

Formal analysis (GC B cell manipulation/FACS analysis): LJG, HFPR, DJH

Formal analysis (Bioinformatics/statistical analysis) MMH, HC, CH, GK, KK, IrM, IlM, ADJ

Interpretation of findings- MMH, SBN, DJH, CT, ADJ

Writing – original draft: PJ, MMH, ADJ

Writing – review & editing: PJ, MMH, WJC, SBN, CT, ADJ

Underlying data verification: MMH, ADJ

