## Supplementary Appendix for "Patterns of oncogene co-expression at single cell resolution in cancer influence survival"

### Investigators

**Authors:** Michal Marek Hoppe, PhD^1^; Patrick Jaynes, PhD^1^; Fan Shuangyi, MD^2^; Yanfen Peng, MSc^1^; Phuong Mai Hoang, PhD^1^; Liu Xin, MA^3^; Sanjay De Mel, MBBS^3,4^; Limei Poon, MBBS^3,4^; Esther Chan, MBBS^3,4^; Joanne Lee, MBBS^3,4^; Ong Choon Kiat, PhD^5^; Tiffany Tang, MBBS^6^; Lim Soon Thye MBBS^6^; Chandramouli Nagarajan, MBBS^7^; Nicholas Francis Grigoropoulos, MBBS, PhD^7^; Soo-Yong Tan, MBBS DPhil^2,4^; Susan Swee-Shan Hue, MBBS^2,4^; Sheng-Tsung Chang, MD^8^, Shih-Sung Chuang, MD^8^; Shaoying Li, MD^9^; Joseph D. Khoury, MD^9^; Hyungwon Choi, PhD^10^; Carl Harris III, PhD^11^; Alessia Bottos, PhD^11^; Laura J. Gay, PhD^12^; Hendrik F. P. Runge, BSc^12^; Ilias Moutsopoulos, BSc^12^; Irina Mohorianu, PhD^12^; Daniel J. Hodson, MBBChir PhD^12^; Pedro Farinha, MD^13^; Anja Mottok, MD^13^; David W. Scott, MBChB PhD^13^; Gayatri Kumar, PhD^14^; Kasthuri Kannan, PhD^14^; Wee-Joo Chng, MBChB PhD^1,4,10^; Yen Lin Chee, MBChB PhD^3,4^; Siok-Bian Ng, MBBS^1,2,4^; Claudio Tripodo, MD PhD^15^; Anand D. Jeyasekharan, MBBS PhD ^1,3,4,10^

**Affiliations:**

^1^Cancer Science Institute of Singapore, National University of Singapore, Singapore

^2^Department of Pathology, Yong Loo Lin School of Medicine, National University of Singapore, Singapore

^3^Department of Haematology-Oncology, National University Health System, Singapore

^4^ NUS Centre for Cancer Research (N2CR), Yong Loo Lin School of Medicine, National University of Singapore (NUS), Singapore

^5^Division of Cellular and Molecular Research, National Cancer Centre Singapore, Singapore

^6^Division of Medical Oncology, National Cancer Centre Singapore, Singapore

^7^Department of Haematology, Singapore General Hospital, Singapore

^8^Department of Pathology, Chi-Mei Medical Center, Tainan City, Taiwan

^9^Department of Hematopathology, Division of Pathology and Laboratory Medicine, The
 University of Texas MD Anderson Cancer Center, Houston, USA ^10^Department of Medicine, Yong Loo Lin School of Medicine, National University of Singapore,
 Singapore

^11^F. Hoffmann-La Roche Ltd, Basel, Switzerland.

^12^Wellcome MRC Cambridge Stem Cell Institute, Cambridge, UK

^13^BC Cancer Research Centre, Vancouver, Canada

^14^Translational Molecular Pathology, The University of Texas MD Anderson Cancer Center, Houston, USA

^15^Tumor Immunology Unit, University of Palermo, Palermo, Italy

### Supplementary Methods

#### Samples and datasets (expanded)

Tonsils (*n*=10) from patients diagnosed with chronic tonsillitis and reactive lymph nodes (*n*=2) from patients diagnosed with reactive follicular hyperplasia collected between 2010 and 2017 at the National University Hospital, Singapore (approved by the Singapore NHG Domain Specific Review Board B study protocol (2015/00176)), comprised a collection of non-malignant tissues to study MYC, BCL2 and BCL6 interaction during normal B-cell development. DLBCL cohorts from NUH (*n*=152), Chi-Mei Medical Center, Taiwan (CMMC cohort, *n*=150), Singapore General Hospital, Singapore (SGH cohort, *n*=67) and from the MD Anderson Cancer Center, USA (MDA cohort *n*=40) were used for quantitative mfIHC analyses in malignant samples. Pre-treatment biopsies of the NUH, SGH and MDA cohorts were used for survival analysis following standard first-line R-CHOP-like (rituximab, cyclophosphamide, doxorubicin hydrochloride, vincristine sulfate, prednisone) therapy. Pre-treatment samples from the British Columbia Cancer Agency (BCA cohort, *n*=274) with first-line R-CHOP-like follow up data were used as a prospective validation cohort ^1^. Full patient characteristics for all above cohorts are provided in Supplementary table10. Relevant ethics approvals from all providing institutions were incorporated into the framework of an NUS IRB approved translational study (H-19-055E). Pre-processed gene expression data along with clinical characteristics was obtained from Gene Expression Omnibus (GEO) for datasets GSE117556 (*n*=928)^2^, GSE125966 (*n*=553)^3^, GSE31312 (*n*=498)^4^, GSE10846 (*n*=420)^5^, GSE32918 (*n*=140)^6^ and GSE87371 (*n*=221)^7^. Raw gene expression and clinical data for Reddy *et al.* (*n*=773) ^8^ and Schmitz *et al.* (*n*=480)^9^ were obtained from the database of Genotypes and Phenotypes (dbGaP). Clinical data associated with the GOYA dataset (GSE125966) was analysed in collaboration with F. Hoffmann-La Roche Ltd.

#### Probabilistic inference of co-localisation

Assuming independent distribution of positivity between MYC, BCL2 and BCL6, a probability-based algorithm using single-marker scores was derived to predict percentage extent of sub-populations i.e. permutations of MYC, BCL2, BCL6-positivity and -negativity, in CD20+ cells in DLBCL samples. For sub-populations involving the lack of expression of a marker, the percentage extent of the negative cell population should be applied, e.g. percentage extent of the MYC+BLC2+BCL6- (M+2+6-) sub-population can be calculated as follows:

$$M+2+6-\%=\left( \frac{MYC\%}{100}\times\frac{BCL2\%}{100} \times\frac{(100-BCL6\%)}{100} \right)\times100\%$$

#### Transforming mRNA expression data into % extent

To transform gene expression data into predicted percentage extent values, the observed single oncogene percentage extent values at the protein level must be similar across different cohorts, coupled with the assumption that there is agreement between mRNA and protein expression ^10^. We observed a strong concordance between the percentage extent of MYC, BCL2 and BCL6 across multiple DLBCL cohorts (Figure4A, Supplementary figure8A, *top*). This allows for a consensus population protein distribution for each oncogene to be established (Supplementary figure8A, *bottom*). To convert quantitative mRNA to inferred single oncogene percentage extent, we first fitted mRNA expression data to the observed percentage extent consensus distribution established from protein expression analysis (Supplementary figure8B-C). The transformation curves were fitted manually using the GSE10846 (Lenz et al.) dataset and validated in the other gene expression datasets (Supplementary figure8C).

For gene expression detected with multiple probes, single probes with best matching distribution characteristics were used. Binary logarithm of mRNA reads was used for transformation; MYC expression data was fitted using the following sigmoid function:

$MYC\%= \frac{1}{1+e^{-0.75\times(MYC-p)}}\times100\%$

where *p* denotes 70^th^ percentile value of MYC expression in the dataset. BCL6 expression data was fitted using the following sigmoid function:

$BCL6\%= \frac{1}{1+e^{-0.6\times(BCL6-p)}}\times100\%$

where *p* denotes 27^th^ percentile value of BCL6 expression in the dataset. Since BCL2 percentage extent follows a binomial distribution in the population, BCL2 mRNA expression values were fitted as follows: mRNA expression values were normalized between the 10^th^ and 100^th^ percentile of values (all values below 10^th^ percentile were assumed to be equal zero) reflecting the approximate prevalence of cases with <1 percentage-positivity extent BCL2 IHC staining. The normalized values (BCL2n) were transformed using the following function:

$BCL2t= \frac{1}{1+{70}^{-(BCL2n/100)}}$

and the BCL2t values were normalized within each cohort into 0-100 percentage extent values.

This workflow for data transformation creates inferred single oncogene percentage extent distributions across GEP cohorts (Supplementary figure6), which can be used in the probabilistic model described in above to estimate the percentage extent of sub-populations (Supplementary table5).

#### Semi-quantitative immunohistochemistry

TMA preparation and staining for MYC, BCL2 and BCL6 for the BCA cohort are described in ref. ^1^ and ref. ^11^. Percentage extent of MYC, BCL2 and BCL6 staining was determined independently by two pathologists. Thresholds for overall tumor positivity for a given marker were: 40% for MYC, 50% for BCL2 and 30% for BCL6. When there was a discrepancy in scoring between the two pathologists to the extent that scores were found to be on opposing sides of a positivity threshold, a consensus score was obtained from a third pathologist.

Machine-learning analysis utilizing the nuclear counterstain image with CD20 membrane boundaries allowed identification of single cells and quantitation of the per-cell intensity of MYC, BCL2, BCL6 and Ki67. From the unmixed images, an intensity-based threshold defined each cell as “positive” or “negative” for the given marker of interest. These thresholds were validated by two independent expert hematopathologists.

#### Correlation of Gene Expression and inferred M+2+6- percentage extent

Gene expression for a consensus of 15314 genes was correlated (Spearman correlation) with inferred M+2+6- percentage extent for seven GEP cohorts. Post-processed gene expression values submitted by the original authors were used of the seven cohorts available at the time of analysis (Sha *et al.*, Reddy *et al.*, McCord *et al.*, Visco *et al.*, Schmitz *et al.*, Lenz *et al.* and Dubois *et al.*). Due to differential formats of post-processed gene expression data, all Spearman rho values for each cohort were standardized to follow similar distributions between cohorts. Genes with an absolute Spearman rho value of > 0.25 and an FDR < 0.001 were considered significant. Genes meeting the significance threshold in 6 out of 7 cohorts were considered hits in this analysis.

#### Differential Gene Expression in Primary GC B-cells

Total RNA was isolated from primary transduced B-cells using TRIzol extraction method. Insert cDNA library creation (250~300bp eukaryotic mRNA) and standard polyA paired-end sequencing on Illumina Hiseq-4000 PE150 was performed by NovogeneAIT. Raw sequencing files were processed using standard pipelines available publicly on the CSI NGS Portal^12^. Gene expression of 18252 genes was compared between four transduced GC B-cell samples of M+2+6+ and four M+2+6- samples from independent donors (see below for details on GC B cell transduction and refs^13,14^). False-discovery rate (FDR) of a two-sided t-test was used to define differently expressed genes. Hits were defined by meeting an arbitrary dynamic threshold criterion defined by the rational function (see dashed line Figure5D):

$$-{log}_{10}({FDR}_{gene})\geq\frac{{{{-log}_{10}(FDR}_{sig})}^{\frac{5}{|FC|}}}{\sqrt{\frac{|FC|}{5}}}$$

Where ${FDR}_{gene}$ is the FDR value of the gene tested, ${FDR}_{sig}$ is an arbitrary threshold of significance of 0.05, and$|FC|$ is the absolute value of ${log}_{2}$ fold change difference between mean expression in M+2+6- and M+2+6+ samples.

#### Single Cell RNA sequencing analysis

For single cell RNA sequencing experiments, primary human GC B cells from a single donor were transduced with BCL2 and BCL6, or BCL2 and MYC (see below for transduction details and refs^13,14^). Seven days after transduction, cells were pooled and subjected to single cell RNA sequencing using the 10X Genomics platform. Fresh transduced GC B cells from the same donor were spiked into the sequencing reaction. Raw fastq files were processed using cellranger (v3.1.0); the alignment was performed against the GRCh38-3.0.0 version of the *Homo sapiens* reference genome; the quantification and filtering of cells were done using default parameters.

Further filtering applied on the expression matrix was based on upper and lower bounds on the distributions of counts and features, and on the proportions of reads incident to mitochondrial DNA (mt%) and ribosomal genes (rp%). Cells with values outside these ranges (counts per cell/sequencing depth > 5000, number of features <2000 or >8000, mt%>15% rp%>50%) were considered outliers and excluded from downstream analyses. Post filtering, mitochondrial and ribosomal genes were excluded from the expression matrix. The expression matrix was log-normalised using the NormalizeData function in the Seurat package (v3.2.2)^15^.

Dimensionality reductions (PCA followed by UMAP), as well as clustering (Louvain method) were conducted in Seurat; the optimal number of clusters was selected based on default clustering parameters. Following an assessment of the stability of clustering results, for the subsequent steps, we focused on the 2000 most abundant genes, determined across all cells in the dataset. Marker genes, determined for each cluster against all other genes, were identified based on differential expression tests (in Seurat) i.e. genes with log_2_(FC) > 0.25, and adjusted p-values, under a Benjamini-Hochberg multiple testing correction, less than 0.05. The data was also made available as a Shiny app^16^.

#### Generation of Immortalized Patient Derived GC B-cells, and CCDN2 analysis

Discarded tonsil tissue was collected after tonsillectomy at Addenbrooke’s ENT Department, Cambridge, UK with written informed consent from the patient’s parent/guardian. Ethical approval for human tissue use was granted by the Health Research Authority Cambridgeshire Research Ethics Committee (REC no. 07/MRE05/44). Human primary germinal centre (GC) B cells were isolated from fresh tonsils with magnetic beads and cultured *in vitro* on irradiated YK6-CD40lg-IL21 follicular dendritic feeder cells in Advanced RPMI-1640 (Invitrogen, cat. no. 12633020) supplemented with 20 % Gibco FCS (Thermo Fisher Scientific, cat. no. 10270-106) and 1 x Gibco Penicillin-Streptomycin-Glutamine (from 100×, Thermo Fisher Scientific, cat. no. 10378016) as previously described ^13,14^. GC B cells were stably transduced with BCL6-T2A-BCL2 and MYC-GFP retrovirus as indicated. Subsequently, cells were stably transduced with either empty vector (EV)-IRES-LyT2 or CCND2-IRES-LyT2 lentivirus. The fraction of EV or CCND2-transduced cells was assessed by flow cytometry after staining for LyT2 with anti-murine CD8a-APC antibody (Miltenyi Biotec, cat. no. 130-117-776) and observed for 30 days.

#### Spatial analysis

The difference in clustering of different oncogene (MYC, BCL2 and BCL6) subpopulations in DLBCL were evaluated for images from the SGH (67 images) and the MDA (87 images) cohorts. We annotated the images and used the point pattern processes to estimate the clustering. In order to identify clustering in different subpopulations we used the Pair-Correlation Function (PCF) ^17,18^.

The PCF is the observed number of pairs of cells that are about *r* units apart, divided by the expected number that would be obtained if the cells were completely random. Mathematically, it is given by,

$$g\left( r \right)= \frac{K'(r)}{2\pi r}$$

where $K(r)$ is the Ripley’s *K*-function that provides an estimate for the expected number of cells within radius *r* of any given cell. This measure can determine if the cells are inhibiting, random or clustering.

First, we annotated the raw images in QuPath by overlaying the cell coordinates using an open source package (opencv2). The annotations were stored as geojson files from which the spatial point pattern objects were generated for each image. For the point pattern objects generated for each cohort, each of the subpopulations were extracted and the clustering was estimated using the PCF function in spatstat ^19^, a comprehensive spatial statistics package. Following this, a distribution of the PCF values for all values of *r* for a specific subpopulation was plotted. The mode around 1 will correspond to the random subset of the cells, while the modes less than and greater than 1 denotes inhibited and clustering subsets, respectively. We performed this analysis for each subpopulation for both SGH and MDA cohorts.

To investigate spatial relationships between different sub-populations, we investigated the sub-population identity of *k*=20 nearest neighbours for each cell (using beforementioned spatial coordinates and an in-house base R script), and compared the percentage extent of such local neighbourhood samples to the overall observed sub-populations extent for a given image. The absolute difference between observed local and global sub-population percentage for each cell we term Δ% and a mean value for all cells of the same sub-population in an image we define as measure of sub-population to sub-population integration or separation.

### Supplementary Figures

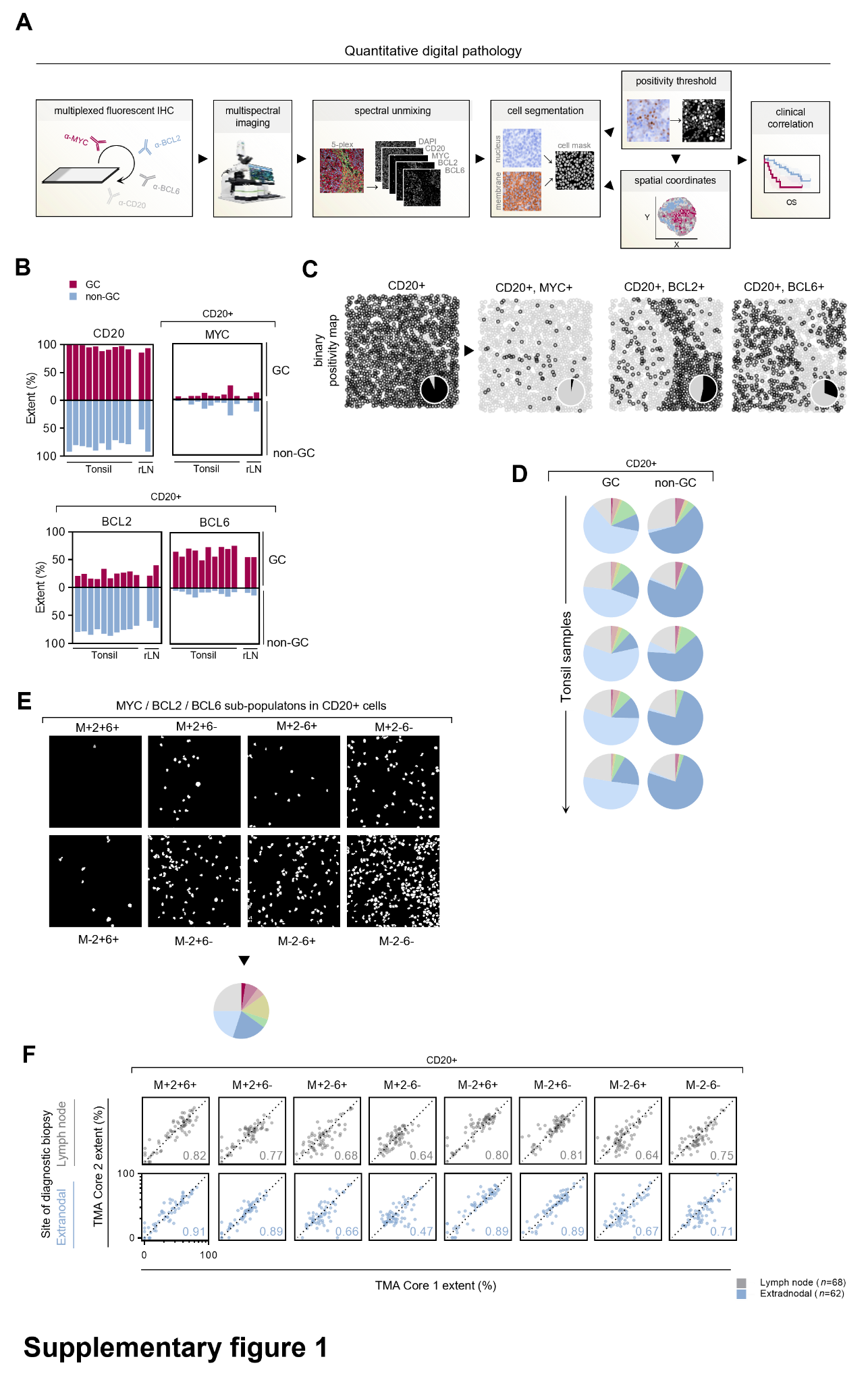

**Supplementary Figure1. Quantitative single-cell analysis of MYC, BCL2 and BCL6 protein expression in B-cells in normal tissues and diffuse large B-cell lymphoma.** A, Schematic workflow of a quantitative digital pathology experiment. B, Quantitation of marker positivity across ten normal tonsil and two normal reactive lymph node samples (rLN). Analysis is spatially resolved between the GC and non-GC zones. C, Spatial map of cellular coordinates based on cell segmentation of images in Figure1A. Marker-positivity is indicated, and a total proportion of positive and negative cells is depicted as a pie chart. D, Five tonsil samples are represented as pie charts for an overview corresponding to Figure1C. E, Positivity masks for sub-populations in example image from Figure1D. F, Correlation of sub-population extent quantification between two biopsies of the same patient for which at least two tissue microarray (TMA) biopsies are available. Correlation is shown separately for lymph node and extranodal biopsies. Spearman rho is indicated for each correlation. Axes are in logarithmic scale and equivalent in all panels.

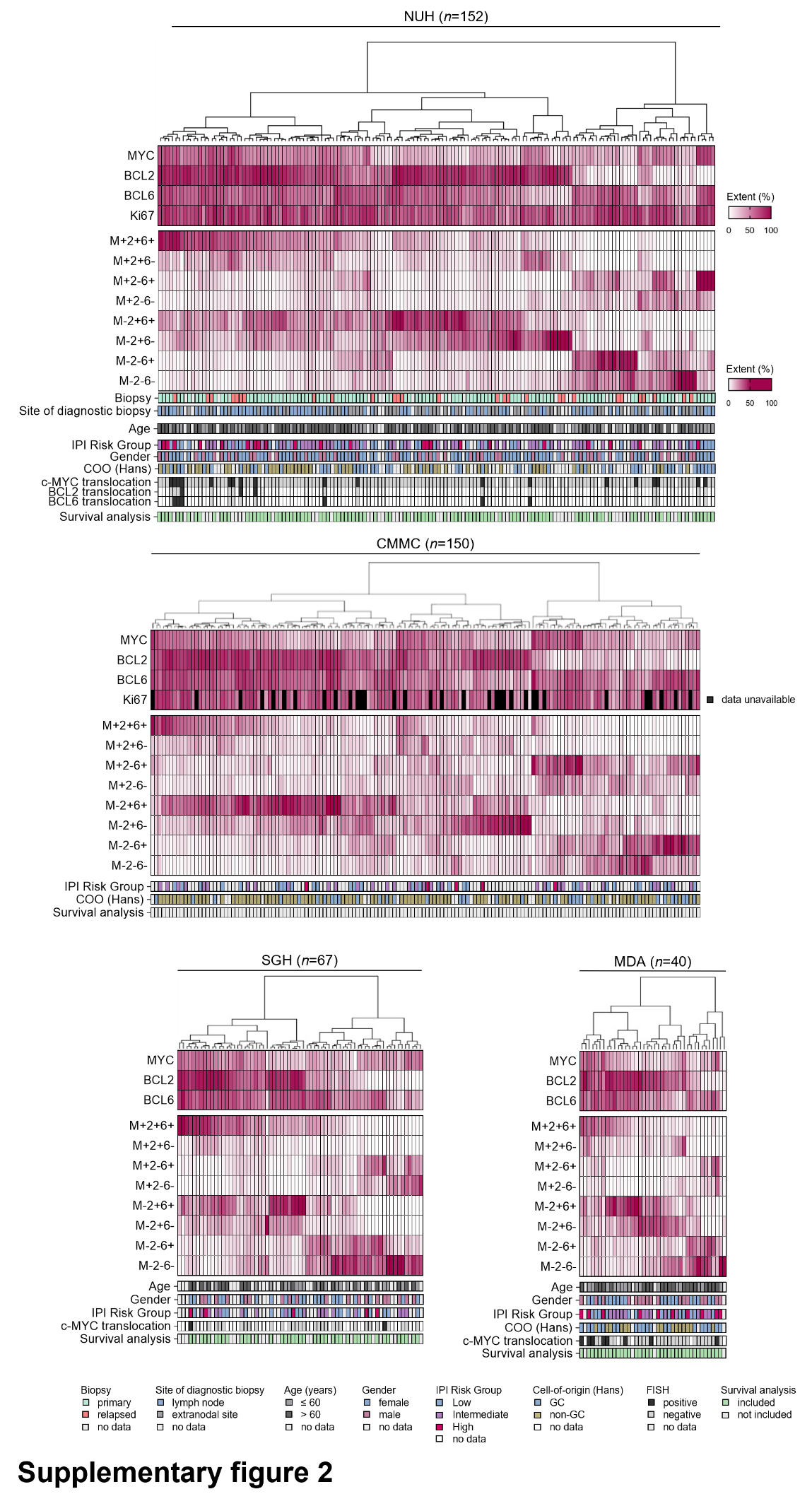

**Supplementary figure2. Global distribution of MYC, BCL2 and BCL6 sub-populations within DLBCL cohorts.** Heat-maps displaying the percentage extent of individual oncogenes and each sub-population within the DLBCL NUH, CMMC, SGH and MDA cohorts. Hierarchical k-means clustering of patients according to sub-population extent is applied. Positivity shading for single oncogenes ranges between 0-100% positivity, whereas shading for sub-populations reflects 0-50% positivity. IPI Risk Group - International Prognostic Index Risk Group, GC - germinal center B-cell like lymphoma. FISH - fluorescence in situ hybridization.

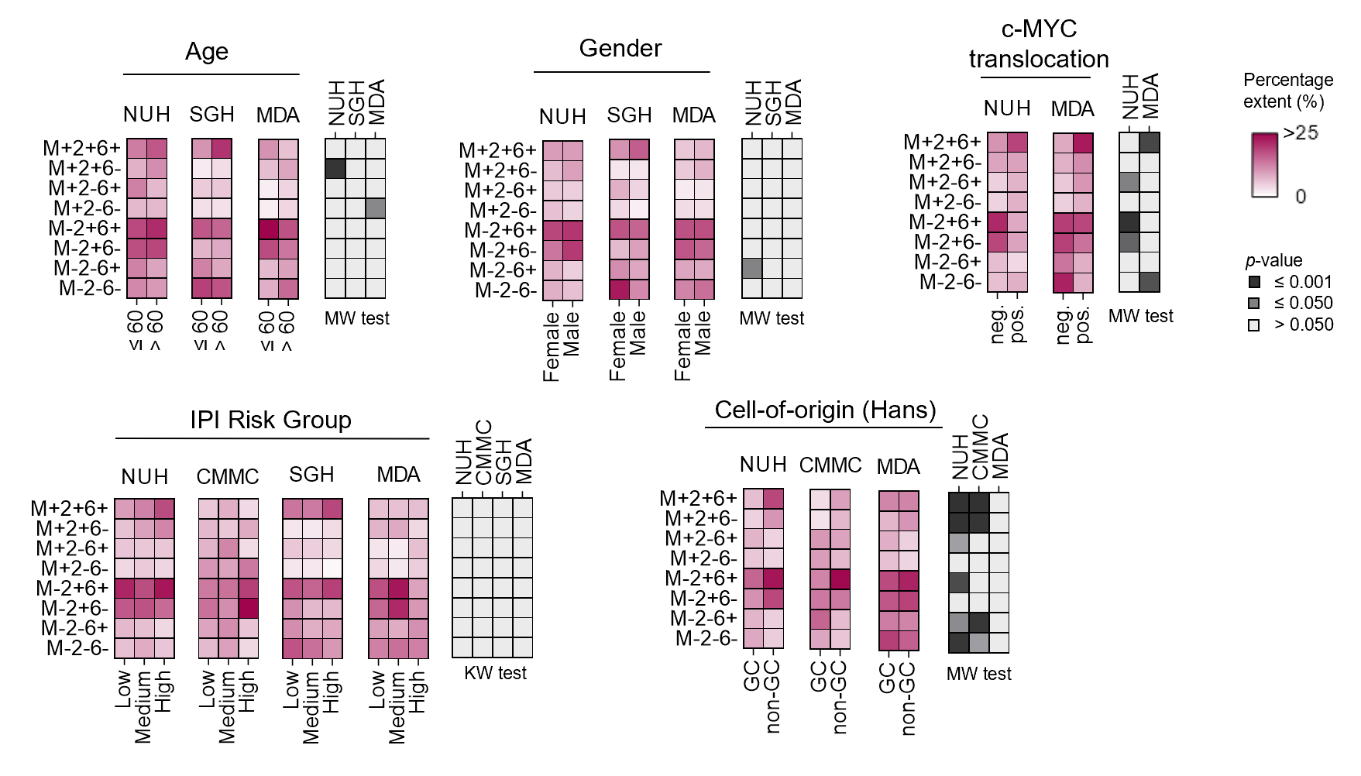

**Supplementary figure3. Graphical correlation of sub-population extent with clinicopathological features.** Mean sub-population extent for each category is shown and statistical test result *p*-value is displayed in the rightmost panel for each variable. MW – Mann-Whitney test, KW - Kruskal–Wallis test..

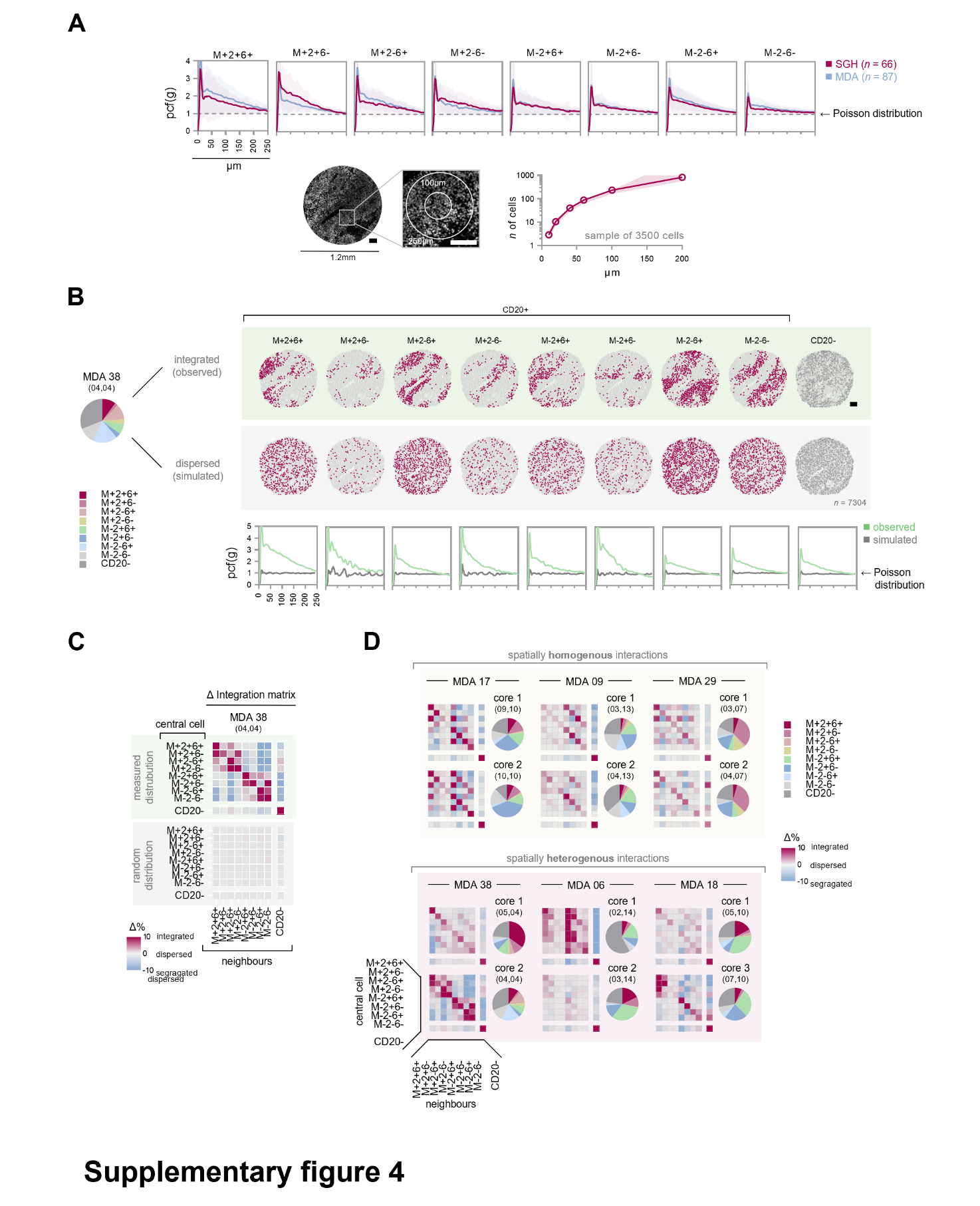

**Supplementary figure4. Spatial heterogeneity of sub-population interactions.** A, Pair correlation function (pcf) for sub-populations to investigate spatial clustering (*top*)*.* Mean results for two independent cohorts (shading is cohort standard deviation). An example tissue microarray core is shown as physical distance reference for spatial analyses (*bottom left*). Absolute number of neighboring cells expected within a given radius (data from 3500 randomly selected cells across all images, mean with standard deviation) (*bottom right*). B, Actual spatial map of sub-populations of an example DLBCL case (*top*). Extent of all sub-populations within the sample is shown on the left. Simulated, hypothetical random distribution of cells for the same case (*middle*). Pair correlation function for the shown sample and its matched simulated random distribution (*bottom*). Scale bars in A and B are 100µm. C, Mean deviations from expected neighbor abundance (Δ%) summarizing cell-cell interactions between sub-populations for the sample shown in B. D, Sub-population interaction matrices from spatially distinct biopsies (cores in tissue microarray) for example DLBCL patients. Biopsies of stable, spatially homogenous, sub-population interaction profiles are grouped (*top*), whereas biopsies of a differing, heterogenous, interaction profile are grouped separately (*bottom*).

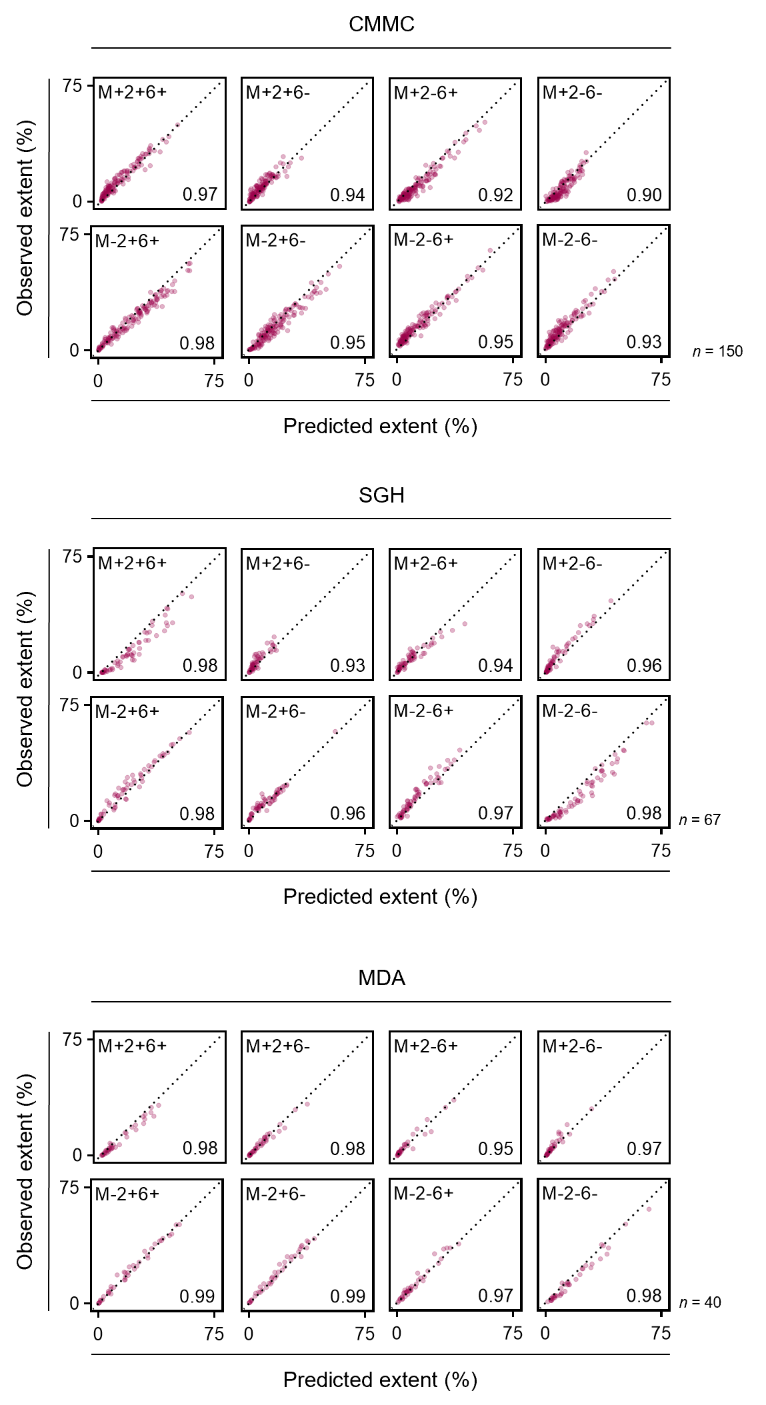

**Supplementary figure5. Correlation of predicted MYC, BCL2 and BCL6 sub-population percentage extent based on single-marker positivity and observed percentage extent in DLBCL cohorts.** Spearman r.

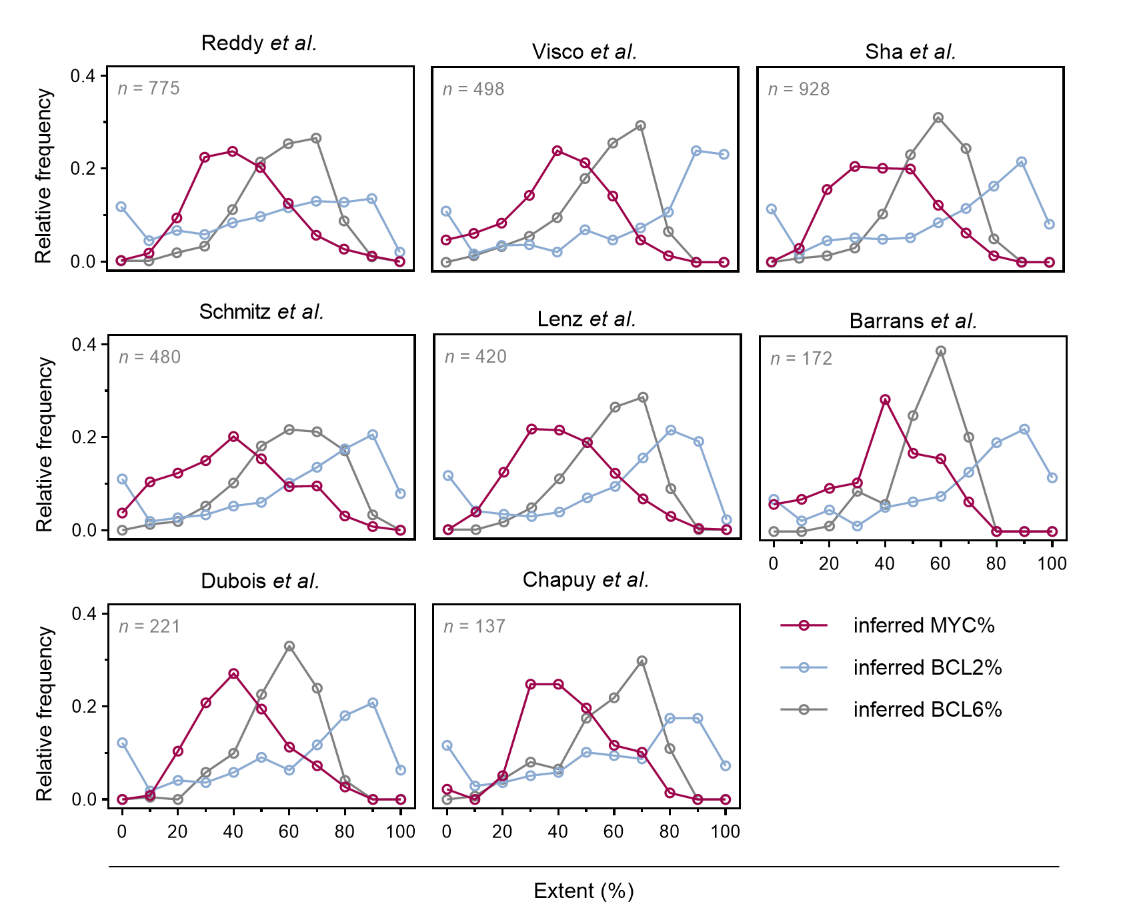

**Supplementary figure6. Distribution of inferred single-marker positivity in GEP cohorts.**

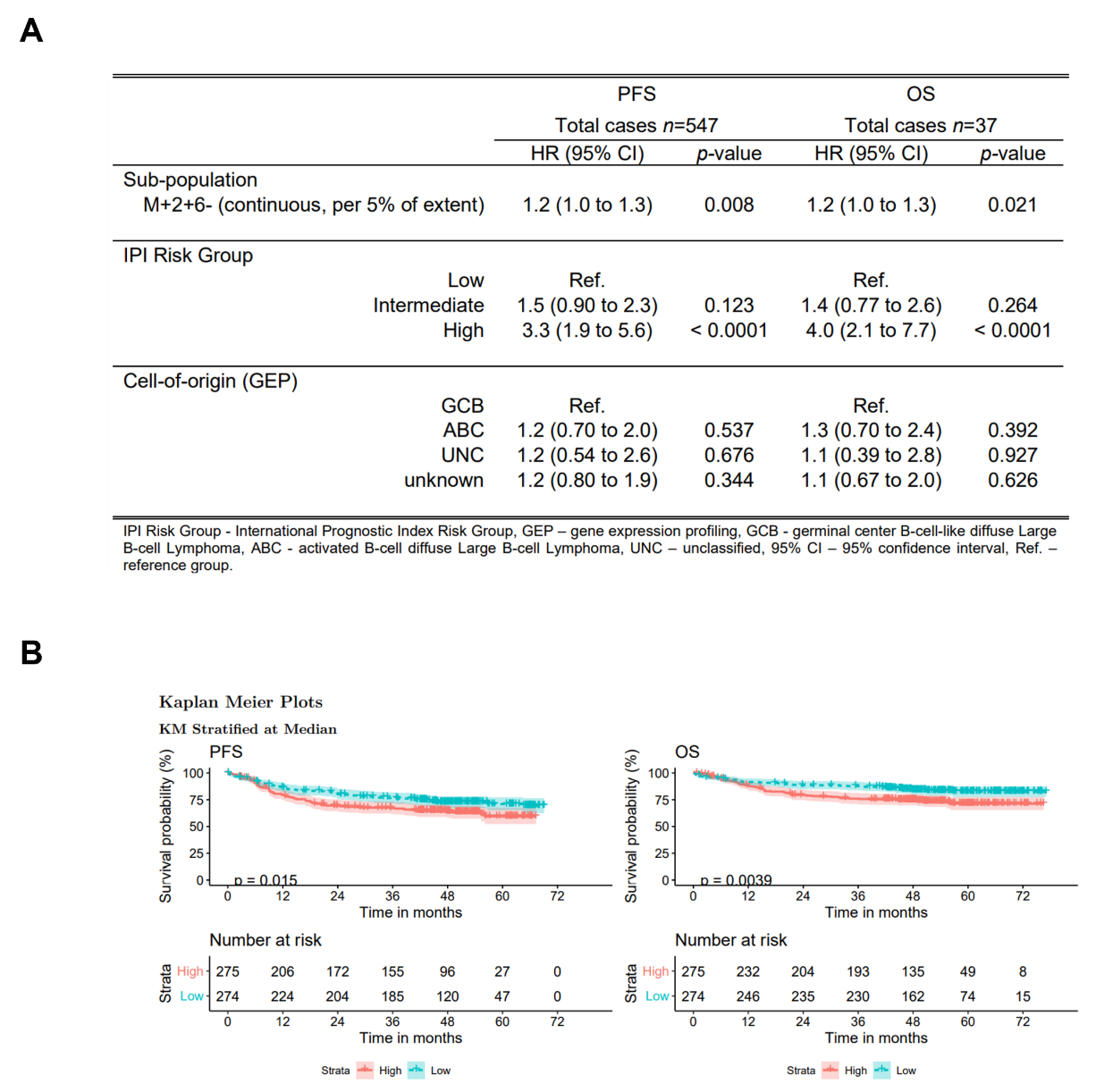

**Supplementary figure7. Independent validation of predictive and prognostic significance of the M+2+6- sub-population in the GOYA clinical trial.** A, Multivariate analysis of continuous M+2+6- percentage extent at 5% increments as a predictor of progression-free survival (PFS) and overall survival (OS) in the GOYA clinical trial (Cox proportional hazards model). B, Kaplan-Meier curves for PFS and OS for patients stratified across the median.

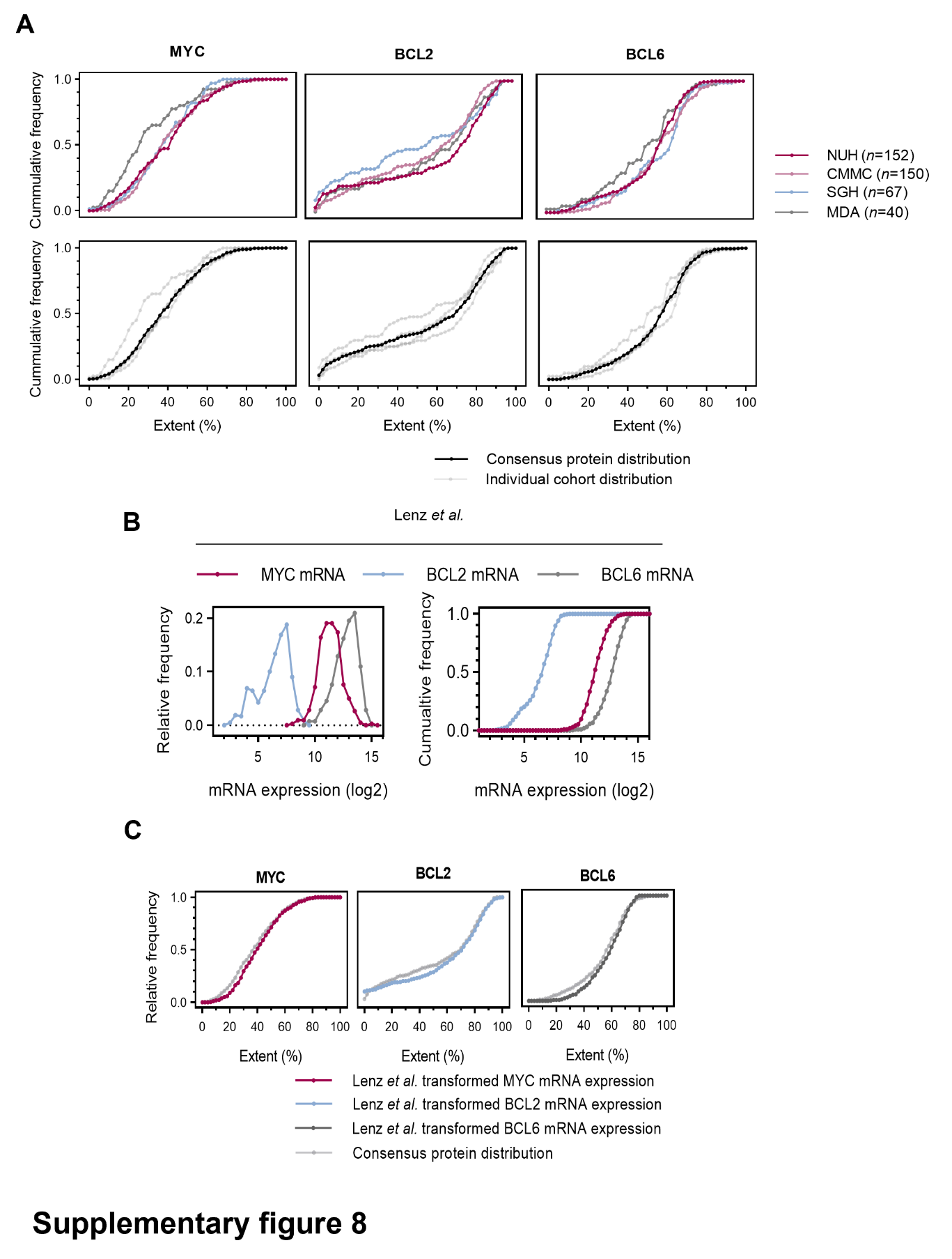

**Supplementary figure8. Transformation of mRNA expression data into percentage extent data**. A, Cumulative histogram of MYC, BCL2 and BCL6 protein percentage extent positivity in DLBCL cohorts (data transformed from Figure4A) (*top*). A consensus single marker cumulative distribution of MYC, BCL2 and BCL6 protein percentage extent positivity across all measured protein cohorts (*bottom*). Inference of percentage extent positivity of MYC, BCL2 and BCL6 single-oncogenes from gene-expression profiling: B, Histograms of mRNA expression of single-oncogenes in the Lenz *et al.* cohort. C, Distribution of transformed mRNA expression of oncogenes in (B) to approximate the measured consensus protein distribution (see Supplementary Methods).

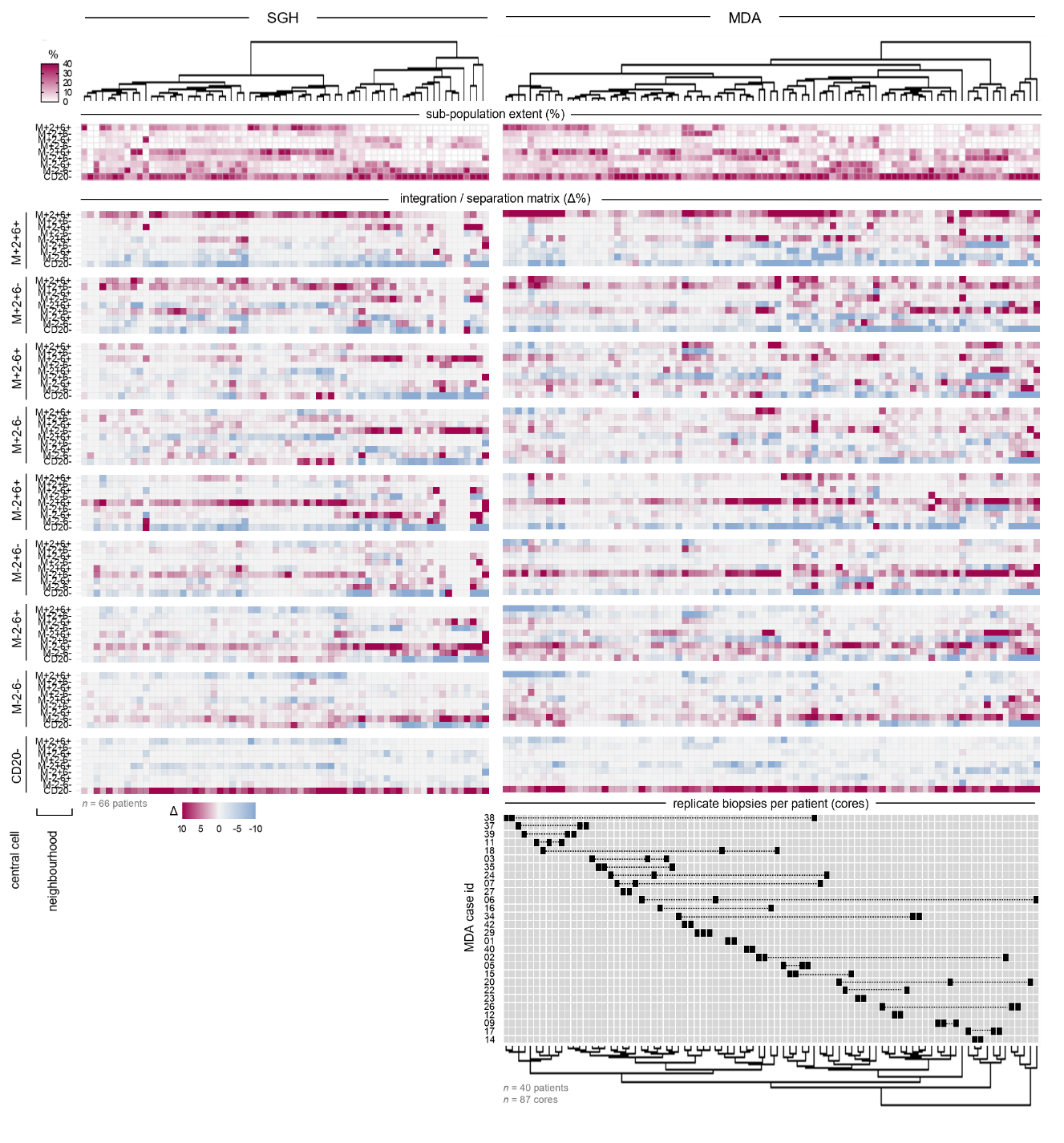

**Supplementary figure9. Global deviations from expected spatial neighbor abundance (Δ%).** Hierarchical clustering (minimum variance method) of measured Δ% for all cases in the SGH and MDA cohorts. Extents of sub-populations are indicated for reference (*top*)*.* For the MDA cohort, multiple biopsies (*n =* 1-3) from the same patient were included in the analysis to determine spatial interaction similarity across spatially distinct regions (*bottom*)*.*

### Supplementary Tables

| Supplementary table1. Manual multiplexed fluorescent immunohistochemistry (mfIHC) staining protocol performed on the NUH and CMMC cohort TMA. | | |
| --- | --- | --- |
| Procedure | Reagent (product; dilution) | Duration (min.) |
| 1^st^ primary antibody incubation | anti-Bcl-6 (Leica, LN22, NCL-564; 1:30) | 60 |
| 1^st^ secondary antibody incubation | anti-Mouse HRP (Dako, K4001) | 10 |
| 1^st^ signal amplification and fluorophore deposition | TSA Opal 670 (PerkinElmer FP1487001KT, 1:100) | 10 |
| 2^nd^ epitope retrieval (antibody stripping) | pH 9 HIER (low power) | 10 |
| 2^nd^ epitope blocking | BSA | 10 |
| 2^nd^ primary antibody incubation | anti-Bcl-2 (Dako, 124, M0887; 1:50) | 60 |
| 2^nd^ secondary antibody incubation | anti-Mouse HRP | 10 |
| 2^nd^ signal amplification and fluorophore deposition | TSA Opal 520 (PerkinElmer FP1487001KT, 1:100) | 10 |
| 3^rd^ epitope retrieval (antibody stripping) | pH 9 HIER (low power) | 10 |
| 3^rd^ epitope blocking | BSA | 10 |
| 3^rd^ primary antibody incubation | anti-c-Myc (Abcam, Y69, ab32072; 1:50) | 30 |
| 3^rd^ secondary antibody incubation | anti-Rabbit HRP (Dako, K4003) | 10 |
| 3^rd^ signal amplification and fluorophore deposition | TSA Opal 570 (PerkinElmer FP1487001KT, 1:100) | 10 |
| 4^th^ epitope retrieval (antibody stripping) | pH 9 HIER (low power) | 10 |
| 4^th^ epitope blocking | BSA | 10 |
| 4^th^ primary antibody incubation | anti-CD20 (DAKO, L26, M0755; 1:2000) | 30 |
| 4^th^ secondary antibody incubation | anti-Mouse HRP | 10 |
| 4^th^ signal amplification and fluorophore deposition | TSA Opal 540 (PerkinElmer FP1487001KT, 1:100) | 10 |
| 5^th^ epitope retrieval (antibody stripping) | pH 9 HIER (high power) | 5 |
| 5^th^ epitope retrieval (continued) | pH 9 HIER (low power) | 10 |
| 5^th^ epitope blocking | BSA | 10 |
| 5^th^ primary antibody incubation | anti-Ki67 (Dako, MIB-1, M7240; 1:50) | 45 |
| 5^th^ secondary antibody incubation | anti-Mouse HRP | 10 |
| 5^th^ signal amplification and fluorophore deposition | TSA Opal 620 (PerkinElmer FP1487001KT, 1:100) | 10 |
| Final antibody stripping | pH 9 HIER (low power) | 10 |
| Counterstaining | DAPI (PerkinElmer FP1490; 1:10 in antibody diluent Dako DKO.S302283) | 5 |
| Mounting | CC mount (Sigma C9368) | 10 |
| NUH – National University Hospital, CMMC - Chi-Mei Medical Center, TMA – tissue microarray, HRP – horseradish peroxidase, TSA – tyramide signal amplification, HIER – heat-induced epitope retrieval, BSA – bovine serum albumin, DAPI – 4′,6-diamidino-2-phenylindole. | | |

| Supplementary table2. Automated multiplexed fluorescent immunohistochemistry (mfIHC) staining protocol performed on the SGH, MDA and BCA cohort TMA. | | |
| --- | --- | --- |
| Procedure | Reagent (product; dilution) | Duration (min.) |
| Bake at 60 °C |  | 20 |
| Dewax | Leica Biosystems Bond Dewax Solution (AR9222) | BondMax default step |
| 1^st^ epitope retrieval (BondMax) | Leica Biosystems Bond Epitope Retrieval Solution 2 (AR9640) | 20 |
| Peroxide Block | BOND Polymer Refine Detection Kit (DS9800) | 10 |
| 1^st^ Primary antibody incubation | anti-c-Myc (Abcam, Y69, ab32072; 1:50) | 20 |
| 1^st^ Post Primary | BOND Polymer Refine Detection Kit (DS9800) | 8 |
| 1^st^ Polymer | BOND Polymer Refine Detection Kit (DS9800) | 8 |
| 1^st^ signal amplification and fluorophore deposition | TSA Opal 520 (PerkinElmer FP1487001KT; 1:100) | 10 |
| 2^nd^ epitope retrieval (BondMax) | Leica Biosystems Bond Epitope Retrieval Solution 1 (AR9961) | 15 |
| 2^nd^ primary antibody incubation | anti-CD20 (DAKO, L26, M0755; 1:1000) | 20 |
| 2^nd^ Post Primary | BOND Polymer Refine Detection Kit (DS9800) | 8 |
| 2^nd^ Polymer | BOND Polymer Refine Detection Kit (DS9800) | 8 |
| 2^nd^ signal amplification and fluorophore deposition | TSA Opal 540 (PerkinElmer FP1494001KT; 1:100) | 10 |
| 3^rd^ epitope retrieval (BondMax) | Leica Biosystems Bond Epitope Retrieval Solution 1 (AR9961) | 20 |
| 3^rd^ primary antibody incubation | anti-Bcl-6 (Leica, LN22, NCL-564; 1:50) | 20 |
| 3^rd^ Post Primary | BOND Polymer Refine Detection Kit (DS9800) | 8 |
| 3^rd^ Polymer | BOND Polymer Refine Detection Kit (DS9800) | 8 |
| 3^rd^ signal amplification and fluorophore deposition | TSA Opal 620 (PerkinElmer FP1495001KT; 1:100) | 10 |
| 4^th^ epitope retrieval (BondMax) | Leica Biosystems Bond Epitope Retrieval Solution 2 (AR9640) | 20 |
| 4^th^ primary antibody incubation | anti-Bcl-2 (Dako, 124, M0887; 1:100) | 20 |
| 4^th^ Post Primary | BOND Polymer Refine Detection Kit (DS9800) | 8 |
| 4^th^ Polymer | BOND Polymer Refine Detection Kit (DS9800) | 8 |
| 4^th^ signal amplification and fluorophore deposition | TSA Opal 690 (PerkinElmer FP1497001KT, 1:100) | 10 |
| Counterstaining | DAPI (PerkinElmer FP1490; 1:10 in antibody diluent Dako DKO.S302283) | 5 |
| Mounting | ProLong Diamond Antifade Mountant, CC mount (Life Technologies) |  |
| SGH – Singapore General Hospital, MDA – MD Anderson Cancer Center, BCA - British Columbia Cancer Agency, TMA – tissue microarray, HRP – horseradish peroxidase, TSA – tyramide signal amplification, HIER – heat-induced epitope retrieval, BSA – bovine serum albumin, DAPI – 4′,6-diamidino-2-phenylindole. | | |

| Supplementary table3. Multivariate analysis of continuous M+2+6- percentage extent at 5% increments as a predictor of overall survival (OS) in the NUH, SGH and MDA cohorts of DLBCL (Cox proportional hazards model). | | | | | | |
| --- | --- | --- | --- | --- | --- | --- |
|  | NUH | | SGH | | MDA | |
|  | Total cases *n*=82  missing values *n*=16 | | Total cases *n*=37  missing values *n*=4 | | Total cases *n*=33  missing values *n*=3 | |
|  | HR (95% CI) | *p-*value | HR (95% CI) | *p-*value | HR (95% CI) | *p-*value |
| Sub-population |  |  |  |  |  |  |
| M+2+6- (continuous, per 5% of extent) | 1.3 (1.1 to 1.7) | 0.011 | 1.6 (1.1 to 2.3) | 0.026 | 1.6 (1.1 to 2.4) | 0.014 |
| IPI Risk Group |  | 0.472 |  | 0.015 |  | 0.224 |
| Low | Ref. |  | Ref. |  | Ref. |  |
| Intermediate | 1.8 (0.69 to 4.8) | 0.230 | 4.4 (0.88 to 21.7) | 0.299 | 1.4 (0.19 to 10.1) | 0.754 |
| High | 1.6 (0.54 to 5.0) | 0.387 | 12.6 (2.3 to 70.1) | 0.005 | 4.6 (0.68 to 31.0) | 0.118 |
| Cell-of-origin (Hans) |  | 0.750 | - | - |  | 0.943 |
| GC | Ref. |  | - | - | Ref. |  |
| non-GC | 1.2 (0.45 to 3.0) |  | - | - | 1.1 (0.19 to 6.0) |  |
| c-MYC translocation status |  | 0.621 | - | - |  | 0.554 |
| negative | Ref. |  | - | - | Ref. |  |
| positive | 0.68 (0.15 to 3.1) |  | - | - | 1.7 (0.31 to 9.1) |  |
| IPI Risk Group - International Prognostic Index Risk Group, GC - Germinal center B-cell-like diffuse Large B-cell Lymphoma, NUH – National University Hospital, SGH – Singapore General Hospital, MDA – MD Anderson Cancer Center, 95% CI – 95% confidence interval, Ref. – reference group. | | | | | | |

| Supplementary table 4. Univariate analysis of clinicopathological features as a predictor of overall survival (OS) after first-line R-CHOP treatment in the NUH, SGH and MDA cohorts of DLBCL (Cox proportional hazards model). | | | | | | |
| --- | --- | --- | --- | --- | --- | --- |
|  | NUH | | SGH | | MDA | |
|  | total cases *n*=90  missing values *n*=8 | | total cases *n*=37  missing values *n*=3 | | total cases *n*=36 | |
|  | HR (95% CI) | *p-*value | HR (95% CI) | *p-*value | HR (95% CI) | *p-*value |
| IPI Risk Group |  | 0.410 |  | 0.019 |  | 0.294 |
| Low | Ref. |  | Ref. |  | Ref. |  |
| Intermediate | 1.5 (0.60 to 3.7) | 0.401 | 6.1 (1.3 to 28.4) | 0.021 | 1.1 (0.22 to 5.5) | 0.914 |
| High | 1.9 (0.73 to 5.2) | 0.184 | 11.3 (2.1 to 61.8) | 0.005 | 2.7 (0.64 to 11.4) | 0.176 |
|  | total cases *n*=85  missing values *n*=13 | | - | | total cases *n*=35  missing values *n*=1 | |
|  | HR (95% CI) | *p-*value | - | - | HR (95% CI) | *p-*value |
| Cell-of-origin (Hans) |  | 0.092 |  | - |  | 0.072 |
| GC | Ref. |  | - | - | Ref. |  |
| non-GC | 2.0 (0.89 to 4.6) |  | - | - | 0.80 (0.23 to 2.8) |  |
|  | total cases *n*=94  missing values *n*=4 | | - | | total cases *n*=34  missing values *n*=2 | |
|  | HR (95% CI) | *p-*value | - | *-* | HR (95% CI) | *p-*value |
| c-MYC translocation status |  | 0.309 |  | - |  | 0.098 |
| negative | Ref. |  | - |  | Ref. |  |
| positive | 0.47 (0.11 to 2.0) |  | - | - | 3.1 (0.81 to 11.5) |  |
| IPI Risk Group - International Prognostic Index Risk Group, GC - Germinal center B-cell-like diffuse Large B-cell Lymphoma, NUH – National University Hospital, SGH – Singapore General Hospital, MDA – MD Anderson Cancer Center, 95% CI – 95% confidence interval, Ref. – reference group. | | | | | | |

Supplementary table5 – in a separate excel file

**Supplementary table5.** Inferred percentage extents of MYC, BCL2, BCL6 and sub-populations in GEP cohorts.

| Supplementary table6. Multivariate analysis of continuous inferred M+2+6- percentage extent at 5% increments as a predictor of overall survival (OS) in cohorts with gene-expression data (Cox proportional hazards model). | | | | | | | | | | | | | | | | |
| --- | --- | --- | --- | --- | --- | --- | --- | --- | --- | --- | --- | --- | --- | --- | --- | --- |
|  | Reddy *et al.* | | Visco *et al.* | | Sha *et al.* | | Schmitz *et al.* | | Lenz *et al.* | | Barrans *et al.* | | Dubois *et al.* | | Chapuy *et al.* | |
|  | total cases *n*=617  missing values *n*=136 | | total cases *n*=424  missing values *n*=46 | | total cases *n*=459  missing values *n*=10 | | total cases *n*=199  missing values *n*=34 | | total cases *n*=164  missing values *n*=69 | | total cases *n*=140 | | total cases *n*=122 | | total cases *n*=97  missing values *n*=4 | |
|  | HR (95% CI) | *p-*value | HR (95% CI) | *p-*value | HR (95% CI) | *p-*value | HR (95% CI) | *p-*value | HR (95% CI) | *p-*value | HR (95% CI) | *p-*value | HR (95% CI) | *p-*value | HR (95% CI) | *p-*value |
| Sub-population |  |  |  |  |  |  |  |  |  |  |  |  |  |  |  |  |
| M+2+6- (continuous, per 5% of extent) | 1.13 (1.05 to 1.22) | 0.002 | 1.12 (1.05 to 1.21) | 0.002 | 1.24 (1.10 to 1.40) | 0.001 | 1.09 (0.94 to 1.25) | 0.272 | 1.16 (1.00 to 1.34) | 0.048 | 1.19 (1.04 to 1.36) | 0.014 | 1.24 (1.06 to 1.45) | 0.009 | 1.27 (1.08 to 1.51) | 0.004 |
| IPI Risk Group |  | <0.001 |  | <0.001 |  | <0.001 |  | <0.001 |  | <0.001 |  | - |  | 0.013 |  | 0.001 |
| Low | Ref. |  | Ref. |  | Ref. |  | Ref. |  | Ref. |  | - | - | Ref. |  | Ref. |  |
| Intermediate | 2.7 (1.8 to 4.2) | <0.001 | 2.8 (1.9 to 4.3) | <0.001 | 1.7 (0.90 to 3.7) | 0.100 | 1.8 (1.1 to 3.2) | 0.027 | 1.6 (0.69 to 3.6) | 0.280 | - | - | 2.6 (0.59 to 11.2) | 0.208 | 4.8 (1.4 to 16.5) | 0.014 |
| High | 5.7 (3.6 to 9.0) | <0.001 | 5.8 (3.6 to 9.5) | <0.001 | 4.1 (2.0 to 8.1) | <0.001 | 5.8 (3.0 to 11.0) | <0.001 | 5.4 (2.2 to 13.3) | <0.001 | - | - | 5.5 (1.3 to 23.7) | 0.023 | 11.8 (3.3 to 42.7) | <0.001 |
| Cell-of-origin (GEP) |  | 0.375 |  | 0.131 |  | 0.955 |  | 0.055 |  | 0.025 |  | 0.019 |  | 0.630 |  | 0.368 |
| GCB | Ref. |  | Ref. |  | Ref. |  | Ref. |  | Ref. |  | Ref. |  | Ref. |  | Ref. |  |
| ABC | 1.7 (0.82 to 1.7) | 0.394 | 1.3 (0.88 to 1.8) | 0.209 | 1.1 (0.63 to 1.9) | 0.762 | 2.1 (1.1 to 4.0) | 0.018 | 3.0 (1.3 to 7.1) | 0.012 | 1.4 (0.79 to 2.5) | 0.214 | 1.5 (0.66 to 3.3) | 0.341 | 0.53 (0.22 to 1.3) | 0.157 |
| UNC | 1.4 (0.88 to 2.1) | 0.168 | 1.7 (0.98 to 3.0) | 0.061 | 1.0 (0.58 to 1.9) | 0.914 | 1.7 (0.90 to 3.3) | 0.099 | 1.2 (0.37 to 4.0) | 0.751 | 0.44 (0.20 to 0.99) | 0.047 | 1.2 (0.43 to 3.3) | 0.737 | <0.001 | 0.968 |
| IPI Risk Group - International Prognostic Index Risk Group,GEP – gene-expression profiling, GCB - Germinal center B-cell-like diffuse Large B-cell lymphoma, ABC - Activated B-cell lymphoma, UNC – unclassified, 95% CI – 95% confidence interval, Ref. – reference group. | | | | | | | | | | | | | | | | |

| Supplementary table7. Multivariate analysis of M+2+6- percentage extent dichotomized at 15% as a predictor of overall survival (OS) in cohorts with gene-expression data (Cox proportional hazards model). | | | | | | | | | | | | | | | | |
| --- | --- | --- | --- | --- | --- | --- | --- | --- | --- | --- | --- | --- | --- | --- | --- | --- |
|  | Reddy *et al.* | | Visco *et al.* | | Sha *et al.* | | Schmitz *et al.* | | Lenz *et al.* | | Barrans *et al.* | | Dubois *et al.* | | Chapuy *et al.* | |
|  | total cases *n*=617  missing values *n*=136 | | total cases *n*=424  missing values *n*=46 | | total cases *n*=459  missing values *n*=10 | | total cases *n*=199  missing values *n*=34 | | total cases *n*=164  missing values *n*=69 | | total cases *n*=140 | | total cases *n*=122 | | total cases *n*=97  missing values *n*=4 | |
|  | HR (95% CI) | *p-*value | HR (95% CI) | *p-*value | HR (95% CI) | *p-*value | HR (95% CI) | *p-*value | HR (95% CI) | *p-*value | HR (95% CI) | *p-*value | HR (95% CI) | *p-*value | HR (95% CI) | *p-*value |
| Sub-population |  | 0.125 |  | 0.010 |  | 0.021 |  | 0.495 |  | 0.044 |  | 0.007 |  | 0.002 |  | 0.039 |
| M+2+6- <15% | Ref. |  | Ref. |  | Ref. |  | Ref. |  | Ref. |  | Ref. |  | Ref. |  | Ref. |  |
| M+2+6- ≥15% | 1.3 (0.93 to 1.9) |  | 1.6 (1.1 to 2.3) |  | 1.8 (1.1 to 2.9) |  | 1.2 (0.69 to 2.2) |  | 2.1 (1.0 to 4.2) |  | 2.2 (1.2 to 3.5) |  | 3.1 (1.5 to 6.4) |  | 2.5 (1.0 to 5.9) |  |
| IPI Risk Group |  | <0.001 |  | <0.001 |  | <0.001 |  | <0.001 |  | <0.001 |  | - |  | 0.008 |  | 0.001 |
| Low | Ref. |  | Ref. |  | Ref. |  | Ref. |  | Ref. |  | - | - | Ref. |  | Ref. |  |
| Intermediate | 2.7 (1.7 to 4.1) | <0.001 | 2.8 (1.9 to 4.3) | <0.001 | 1.8 (0.92 to 3.3) | 0.088 | 1.8 (1.1 to 3.2) | 0.028 | 1.3 (0.56 to 3.0) | 0.559 | - | - | 2.4 (0.56 to 10.7) | 0.233 | 4.6 (1.3 to 16.0) | 0.016 |
| High | 5.4 (3.4 to 8.6) | <0.001 | 5.7 (3.5 to 9.2) | <0.001 | 4.2 (2.1 to 8.3) | <0.001 | 5.6 (2.9 to 10.8) | <0.001 | 5.2 (2.1 to 12.7) | <0.001 | - | - | 5.6 (1.3 to 24.1) | 0.020 | 11.5 (3.2 to 41.7) | <0.001 |
| Cell-of-origin (GEP) |  | 0.191 |  | 0.107 |  | 0.792 |  | 0.009 |  | 0.031 |  | 0.037 |  | 0.858 |  | 0.582 |
| GCB | Ref. |  | Ref. |  | Ref. |  | Ref. |  | Ref. |  | Ref. |  | Ref. |  | Ref. |  |
| ABC | 1.3 (0.91 to 1.9) | 0.143 | 1.3 (0.92 to 1.9) | 0.131 | 1.2 (0.70 to 2.1) | 0.495 | 2.4 (1.4 to 4.3) | 0.003 | 3.0 (1.3 to 7.1) | 0.013 | 0.78 (0.43 to 1.4) | 0.413 | 1.2 (0.55 to 2.8) | 0.604 | 0.63 (0.26 to 1.5) | 0.298 |
| UNC | 1.4 (0.93 to 2.2) | 0.105 | 1.7 (0.97 to 3.0) | 0.063 | 1.1 (0.60 to 1.9) | 0.810 | 1.9 (1.0 to 3.5) | 0.047 | 1.2 (0.38 to 4.1) | 0.715 | 0.35 (0.15 to 0.78) | 0.010 | 1.2 (0.45 to 3.4) | 0.684 | <0.001 | 0.967 |
| IPI Risk Group - International Prognostic Index Risk Group,GEP – gene-expression profiling, GCB - Germinal center B-cell-like diffuse Large B-cell lymphoma, ABC - Activated B-cell lymphoma, UNC – unclassified, 95% CI – 95% confidence interval, Ref. – reference group. | | | | | | | | | | | | | | | | |

Supplementary table8 – in a separate excel file

**Supplementary table8**. Correlation of inferred M+2+6- percentage extent with gene-expression in GEP cohorts.

Supplementary table9 – in a separate excel file

**Supplementary table9**. Differential gene expression analysis of primary germinal center (GC) B-cells with M+2+ and M+2+6+ overexpression.

| Supplementary table10 . Clinicopathologic characteristics of DLBCL patients evaluated by multiplexed fluorescent immunohistochemistry (mfIHC) in this study. | | | | | | | | | | |
| --- | --- | --- | --- | --- | --- | --- | --- | --- | --- | --- |
| Cohort | NUH | | CMMC | | SGH | | MDA | | BCA | |
|  | Total cases | Survival analysis | Total cases | Survival analysis | Total cases | Survival analysis | Total cases | Survival analysis | Total cases | Survival analysis |
|  | *n* (%) | *n* (%) | *n* (%) | *n* (%) | *n* (%) | *n* (%) | *n* (%) | *n* (%) | *n* (%) | *n* (%) |
| Number of patients | 152 (100%) | 98  (100%) | 150 (100%) | 0 | 67  (100%) | 41  (100%) | 40  (100%) | 36  (100%) | 303  (100%) | 274  (100%) |
| Age |  |  |  |  |  |  |  |  |  |  |
| ≤60 | 59 (38.8%) | 40 (40.8%) | 0 (0%) | - | 20 (29.9%) | 20 (48.8%) | 16 (40.0%) | 16 (44.4%) | 114 (37.6%) | 111 (40.5%) |
| >60 | 70 (46.1%) | 58 (59.2%) | 0 (0%) | - | 21 (31.3%) | 21 (51.2%) | 20 (50.0%) | 20 (55.6%) | 165 (54.5%) | 163 (59.5%) |
| no data | 23 (15.1%) | - | 150 (100%) | - | 26 (38.8%) | - | 4 (10.0%) | - | 24 (7.9%) | - |
| Gender |  |  |  |  |  |  |  |  |  |  |
| Female | 46 (30.3%) | 35 (35.7%) | 0 (0%) | - | 19 (28.4%) | 19 (46.3%) | 17 (42.5%) | 17 (47.2%) | 100 (33.0%) | 100 (36.5%) |
| Male | 83 (54.6%) | 63 (64.3%) | 0 (0%) | - | 22 (32.8%) | 22 (53.7%) | 19 (47.5%) | 19 (52.8%) | 174 (57.4%) | 174 (63.5%) |
| no data | 23 (15.1%) | - | 150 (100%) | - | 26 (38.8%) | - | 4 (10.0%) | - | 29 (9.6%) | - |
| IPI Risk Group |  |  |  |  |  |  |  |  |  |  |
| Low | 38 (25.0%) | 31 (31.6%) | 31 (20.7%) | - | 17 (25.4%) | 17 (41.5%) | 13 (32.5%) | 13 (36.1%) | 90 (29.7%) | 87 (31.8%) |
| Intermediate | 53 (34.9%) | 39 (39.8%) | 31 (20.7%) | - | 16 (23.9%) | 16 (39.0%) | 13 (32.5%) | 13 (36.1%) | 118 (38.9%) | 117 (42.7%) |
| High | 22 (14.5%) | 20 (20.4%) | 5 (3.3%) | - | 5 (7.5%) | 5 (12.2%) | 10 (25.0%) | 10 (27.8%) | 47 (15.4%) | 46 (16.8%) |
| no data | 39 (25.6%) | 8 (8.2%) | 83 (55.3%) | - | 29 (43.3%) | 3 (7.3%) | 4 (10.0%) | - | 48 (15.8%) | 24 (8.8%) |
| Cell-of-origin (Hans) |  |  |  |  |  |  |  |  |  |  |
| GC | 48 (31.6%) | 42 (42.9%) | 36 (24.0%) | - | 0 (0%) | 0 (0%) | 20 (50.0%) | 20 (55.5%) | 166 (54.8%) | 164 (59.9%) |
| non-GC | 48 (31.6%) | 43(43.9%) | 91 (60.7%) | - | 0 (0%) | 0 (0%) | 15 (37.5%) | 15 (41.7%) | 110 (36.3%) | 109 (39.8%) |
| no data | 56 (36.8%) | 13 (13.3%) | 23 (15.3%) | - | 67 (100%) | 41 (100%) | 5 (12.5%) | 1 (2.8%) | 27 (8.9%) | 1 (0.4%) |
| c-MYC translocation status |  |  |  |  |  |  |  |  |  |  |
| Negative | 124 (81.6%) | 81 (82.7%) | 0 (0%) | - | 31 (46.3%) | not included (little positive cases) | 26 (65.0%) | 26 (72.2%) | 228 (75.2%) | 228 (83.2%) |
| Positive | 19 (12.5%) | 13 (13.3%) | 0 (0%) | - | 2 (3.0%) |  | 8 (20.0%) | 8 (22.2%) | 41 (13.5%) | 41 (15.0%) |
| no data | 9 (5.9%) | 4 (4.1%) | 150 (100%) | - | 34 (50.7%) |  | 6 (15.0%) | 2 (5.6%) | 34 (11.2%) | 5 (1.8%) |
| BCL2 translocation status |  |  |  |  |  |  |  |  |  |  |
| Negative | 16 (10.5%) | 11 (11.2%) | 0 (0%) | - | - | - | - | - | 175 (57.8%) | 175 (63.9%) |
| Positive | 3 (2.0%) | 2 (2.1%) | 0 (0%) | - | - | - | - | - | 77 (25.4%) | 77 (28.1%) |
| no data | 133 (87.5%) | 85 (86.7%) | 150 (100%) | - | - | - | - | - | 51 (16.8%) | 22 (8.0%) |
| BCL6 translocation status |  |  |  |  |  |  |  |  |  |  |
| Negative | 13 (8.6%) | 8 (8.2%) | 0 (0%) | - | - | - | - | - | 203 (67.0%) | 203 (74.1%) |
| Positive | 6 (3.9%) | 5 (5.1%) | 0 (0%) | - | - | - | - | - | 55 (18.2%) | 55 (20.1%) |
| no data | 133 (87.5%) | 85 (86.7%) | 150 (100%) | - | - | - | - | - | 45 (14.9%) | 16 (5.8%) |
| Biopsy |  |  |  |  |  |  |  |  |  |  |
| Primary | 104 (68.4%) | 98 (100%) | - | - | 67 (100%) | 41 (100%) | 40 (100%) | 36 (100%) | 303 (100%) | 274 (100%) |
| Relapsed | 28 (18.4%) | - | - | - | - | - | - | - | - | - |
| no data | 20 (13.2%) | - | 150 (100%) | - | - | - | - | - | - | - |
| IPI Risk Group - International Prognostic Index Risk Group, GC - Germinal center B-cell-like diffuse Large B-cell Lymphoma, NUH – National University Hospital, CMMC - Chi-Mei Medical Center, SGH – Singapore General Hospital, MDA – MD Anderson Cancer Center, BCA - British Columbia Cancer Agency. | | | | | | | | | | |

### Supplementary Results

#### Spatial interaction of oncogenic B-cell sub-populations is clustered and non-random

Single-cell resolved image data with spatial coordinates enables the assessment of the interaction patterns of the eight MYC, BCL2, BCL6 sub-populations. Analysing spatial sub-population data of the SGH and MDA cohorts, we first applied a pair correlation function (PCF)^17,18^ to investigate whether each sub-population tends to cluster or rather shows a scattered spatial pattern. A PCF represents the ratio between observed pairs of cells within a given boundary and expected pairs of cells within a given boundary if the cells were completely random. The PCF graphs in Supplementary figure4A demonstrate that each subpopulation deviates from random (Poisson) spatial patterning (PCF= 1) within a 250µm radius of a given cell. Specifically, for each subpopulation, at small distances we see a sharp increase in the PCF that tapers off as the distance increases, indicative of a clustered cell pattern. This is true for both the SGH and MDA cohorts. Supplementary figure4B illustrates this visually for a single patient: each sub-population shows a tendency to group in space within the tissue and does not display a random spatial Poisson distribution, the latter of which has been generated by random simulation and depicted in Supplementary figure4B, *middle*.

To further quantify spatial heterogeneity between sub-populations, we calculated for each cell the percentage % deviation (Δ%) of the observed from the expected sub-population extent within the cell’s local neighbourhood (20 cells). In other words, if cells were distributed randomly in space, the observed abundance of a particular sub-population in the neighbourhood of any given cell would match the overall sub-population extents measured for a tumor. However, if an over- or under-representation of a particular sub-population occurs in topological neighbourhood of a given cell, this deviation provides a quantitative depiction of local interactions for that cell. Supplementary figure4C demonstrates that each subpopulation (defined here by MYC, BCL2 and BCL6) has a unique pattern in terms of the range of Δ% scores of other subpopulations within their local neighbourhood. While cell groups randomly distributed in space (simulated) are completely dispersed and show no interaction (Supplementary figure4C, *bottom*), empirical measurement suggests that cells of a particular sub-population cluster with the same cell type (as shown in Supplementary figure4A) but also of different types and thus show an integrated interaction with one another (Supplementary figure4C, *top*, e.g. M+2+6- with M+2-6-). Alternatively, cells of two sub-types may spatially not co-occur, and therefore show a segregated interaction (e.g. M+2+6+ with M-2-6+ in the example tumor sample, Supplementary Figure4C, *top*). Such interactions can only be empirically established through spatial investigation, and provide a novel and independent characteristic that is patient-specific (Supplementary figure9). These interaction patterns can be stable across different regions of the tumor for the same patient (Supplementary figure4D, *top*, see Supplementary figure9, clustering of MDA cohort with multiple cores per patient), or heterogenous with spatially varying interaction patterns in different cores from the same patient (Supplementary figure4D, *bottom*). We conclude from these investigations that B-cell sub-populations of different oncogenic co-expression aggregate spatially in a non-random manner (likely reflecting clustering due to parent cell - daughter cell relationships or, alternatively, location of a local stimulates arising from the microenvironment such as T follicular helper/ follicular dendritic cells) and show spatial interactions that are patient-specific.
